# Genome restructuring and lineage diversification of *Cryptococcus neoformans* during chronic infection of human hosts

**DOI:** 10.1101/2025.02.17.25320472

**Authors:** Marhiah C. Montoya, Kayla Wilhoit, Debra Murray, John R. Perfect, Paul M. Magwene

## Abstract

Classified as a critical public health threat by the World Health Organization, *Cryptococcus neoformans* infections with significant morbidity and mortality. Reports of cryptococcosis persistence, relapse, and reinfection date back to the 1950s, yet the factors driving chronic infections remain poorly understood. A major challenge is the scarcity of serial patient specimens and detailed medical records to study the simultaneous evolution of the pathogen and host health status. This study provides the first genomic and phenotypic analysis of in-host evolution of *C. neoformans* during chronic infections lasting over a year in six immunocompromised patients. We find fungal genome evolution during persistent infection is characterized by large-scale genome restructuring and increasing genomic heterogeneity. Phenotypic changes show diversification in virulence traits and antifungal susceptibility. Genotypically and phenotypically distinct sub-lineages arise and co-persist within the same tissues, consistent with a model of diversifying selection and niche partitioning in the complex environment of human hosts.

## 2 Introduction

*Cryptococcus neoformans* is a basidiomycete yeast that causes invasive pulmonary and cerebral fungal infections in humans; such infections are associated with both high morbidity and mortality (Rajasingham et al., 2022). Most cases of cryptococcal meningitis infection are either resolved with antifungal therapy or fatal within a time scale of months (Dao et al., 2024; Rajasingham et al., 2017) but clinical cases of long-term cryptococcosis persistence, relapse, and reinfection have been well documented since the 1950s (Benn et al., 1984; Blasi et al., 2001; Haynes et al., 1995; Lan et al., 2001; Sullivan et al., 1996; White & Arany, 1958). Data regarding the prevalence and presentation of chronic cryptococcal infections beyond the standard 10-week treatment period are scarce but, compared to acute resolved infections, chronic cryptococcal meningoencephalitis often leads to worse outcomes including higher morbidity, mortality, and a myriad of neurological issues (Mishra et al., 2018; Nascimento et al., 2016). The persistence of high morbidity and mortality rates associated with chronic cryptococcal infections, even in the presence of advanced healthcare resources, highlights the pressing need to identify key factors, on both the host and pathogen side of the equation, that contribute to chronic cryptococcal infections and influence health outcomes. The pathogenic components contributing to chronic cryptococcosis remain unclear, but existing data suggest a link to specific fungal (Blasi et al., 2001; Boyce et al., 2017; Brandt et al., 2001; Chen et al., 2017a; Fernandes et al., 2018; Mondon et al., 1999; Montoya et al., 2021; Nascimento et al., 2016; Pfaller et al., 1998; Rhodes, Beale, et al., 2017; Yamazumi et al., 2003) or human (Bratton et al., 2012; Nascimento et al., 2016; Salazar et al., 2020) factors that include fungal genotypes, virulence phenotypes, in-host microevolution, underlying host disease, inadequate antifungal therapy, and missed or delayed diagnoses.

Here we provide one of the most in-depth genomic and phenotypic analyses to date of how *C. neoformans* lineages evolve in human hosts during chronic infections. We find that even from just a single hospital system in the southeastern US, chronic cryptococcal infections can be caused by a broad range of molecular types, including genetically diverse hybrid lineages. Infective strains can be either haploid or diploid, and genome evolution during chronic infection is characterized by extensive genomic remodeling involving changes such as aneuploidy, large scale duplications and deletions, and loss-of-heterozygosity. We detect patterns of branching evolution with multiple genotypically distinct sub-lineages co-occurring in single patient samples and persisting over time. The high level of within-patient genomic variation we observed is accompanied by extensive within-patient diversification and heterogeneity of *Cryptococcus* virulence phenotypes such as capsule size, melanization, and cell shape as well as decreased susceptibility to antifungal drugs such as amphotericin B and fluconazole. In sum, the patterns of genomic and phenotypic evolution we observe during chronic cryptococcal infection are consistent with strong diversifying selection, likely driven by adaptation to both the complex niches present in human hosts as well as the therapeutic interventions that patients under clinical care receive.

## 3 Results

### 3.1 Patient cohort characteristics, morbidity, and clinical outcomes

We studied cases of chronic cryptococcal infection treated in the Duke University Health System (DUHS) between January 1, 1991, and December 31, 2022. Chronic cryptococcosis was defined as cases in which patients had two or more *Cryptococcus*-positive samples, separated by at least 365 days, with detailed care and treatment information available in electronic medical records. Six patients in our study population met these criteria. The average age of these six patients was 36.1 years (age range 29–56 years). Five of the patients were HIV-positive non-Hispanic Black biological males (NC1, NC6, NC8, NC83, NC94) and the sixth was a non-Hispanic White biological female (NC183) who had a bilateral lung solid organ transplant (Figure 1). Patients had between two and thirteen serial specimens available for analysis (Table S1). All HIV-positive patients experienced cryptococcal meningitis (CM), during which cerebrospinal fluid (CSF) specimens were collected. Additionally, patient NC1 had a blood specimen (incident isolate unavailable) and a sputum specimen available prior to dissemination into the CNS. Patient NC94 had a blood specimen taken at their incident encounter prior to fungal dissemination into the CNS, then a blood and CSF specimen at their subsequent encounter. Patient NC183’s infection and specimens were localized to the lungs.

**Figure 1:**
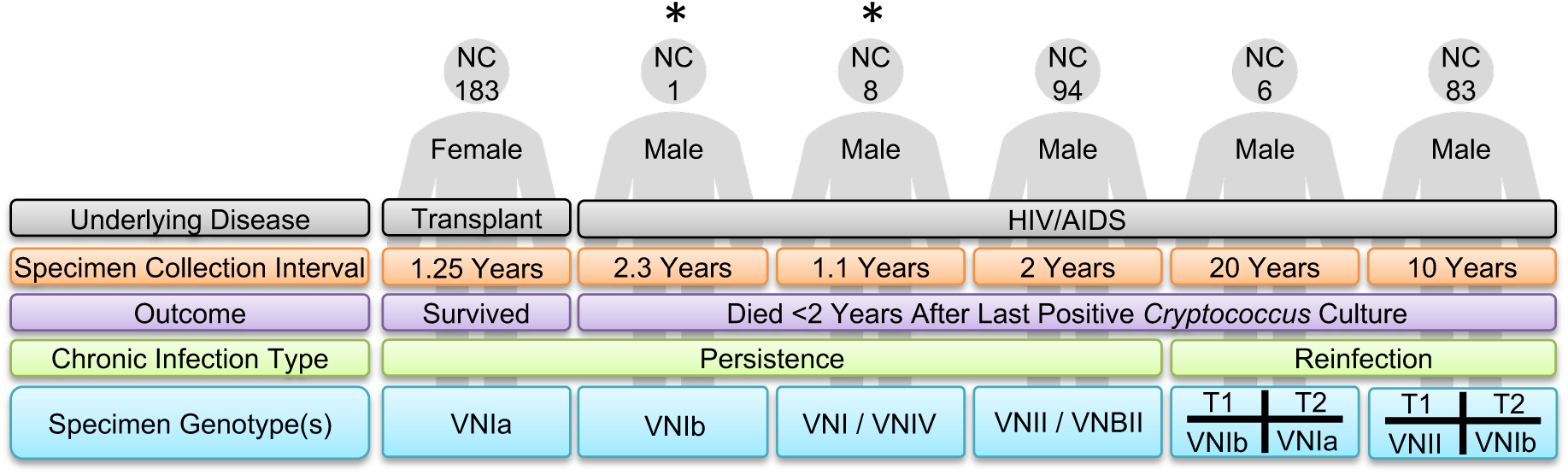
Clinical characteristics of the chronic cryptococcosis patient cohort. Within the DUHS clinical lab mycology collection, six patients were identified as having at least two specimens collected greater than one year apart. Asterisks indicate patients that died while admitted for active cryptococcal meningoencephalitis.

Throughout their chronic cryptococcal infections, patients NC1, NC6, NC8, and NC83 experienced progressive cryptococcosis-related morbidities. Specifically, patient NC1 initially had a normal neurological exam but over the course of their ∼2.5 year CM infection they became consistently non-focal and ultimately deteriorated to complete unresponsiveness. Patient NC6 experienced aphasia that, despite speech therapy, did not return to normal and developed a recurrent seizure disorder during a CM-related episode of acute encephalopathy. Patient NC8 became blind, developed hearing loss with complete deafness in one ear, lost normal speech ability, and had a severe reduction in cognitive ability. Patient NC83 experienced vision decline and was classified as disabled without any neurological deficits. Patient NC94 did not have any CM-specific morbidities, likely attributable to successful antifungal therapy.

NC1 and NC8 died while admitted for acute CM infections 8 days and 48 days after their last *Cryptococcus*-positive CSF culture, respectively. NC6 and NC94 died 248 days and 374 days after their last *Cryptococcus*-positive CSF culture, respectively, due to deteriorating health, organ failure related to HIV infection, and non-*Cryptococcus* co-infections. Lastly, NC83 died from a traumatic injury 750 days after their last *Cryptococcus*-positive CSF culture. NC183 was the only patient included in this study who survived for more than two years after their last positive cryptococcal culture.

### 3.2 Patient isolates exhibit both inter- and intra-specimen phenotypic heterogeneity

Since the 1990s the DUHS Clinical Microbiology Laboratory intermittently collected *Cryptococcus*-positive patient specimens as part of routine clinical care and not according to a research-specific standardized procedure.

Since no standard purification had been carried out in the clinical labs, the first step in specimen analysis consisted of purification of microbial cultures and phenotypic analysis to ensure the presence of *Cryptococcus* and exclude any non-*Cryptococcus* co-infecting organisms. This process included screening individual yeast colonies from each patient specimen using a variety of culturing conditions. Each patient specimen was cultured on nutrient-rich media and nutrient-limiting melanin induction media under standard culturing (ambient air, 30°C) or host-like stress conditions (5% CO_2_, 37°C). In addition, India ink staining and microscopy were used to evaluate cell morphology and to confirm the presence of a polysaccharide capsule.

Patient specimens that showed phenotypic variation under any single condition underwent colony purification and multiple colonies were isolated (Figure S1) and re-phenotyped (Figures S2-S5). Comparing colony phenotypes under multiple growth conditions revealed phenotypically distinct sub-populations within many patient specimens (i.e., intra-specimen heterogeneity) (Figures 2A, S6) and between serial specimens from the same patient (i.e., inter-specimen heterogeneity) (Figures 2B, S6). The stability of colony-specific phenotypes was confirmed by conducting subsequent phenotypic assessments on each distinct colony (hereafter referred to as a strain). While it is commonly understood that phenotypic heterogeneity can exist within a genotypically homogenous population, the significant divergence in colony phenotypes prompted us to conduct whole genome sequencing to determine if phenotypically distinct colonies represented genotypically distinct cryptococcal strains.

**Figure 2:**
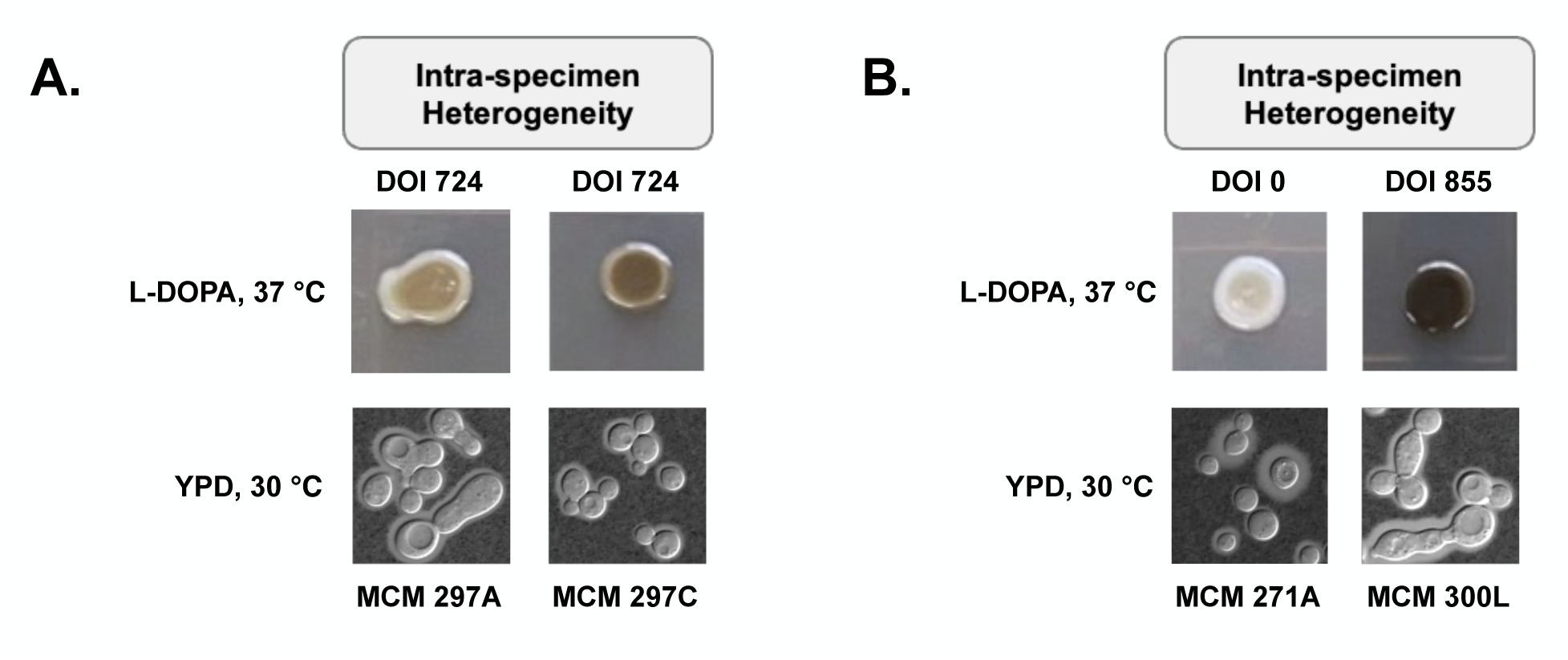
Phenotypic heterogeneity in melanization and cell shape is observed within single patient specimens and between serial specimens from the same patient. A) Intra-specimen phenotypic heterogeneity between two isolates from a CSF specimen collected on DOI 724. B) Inter-specimen heterogeneity between isolates collected at DOI 0 (sputum) and DOI 855 (CSF).

### 3.3 Genome sequencing and lineage assignment

After specimen purification, the genomes of each phenotypically distinct strain was sequenced on the Illumina NovaSeq platform, using 150bp paired-end reads. To identify appropriate species and lineage assignments for further analysis, sequencing reads were initially aligned to a concatenated reference genome consisting of whole genome assemblies of representative strains from each of the major *Cryptococcus* species (*C. neoformans*, *C. deneoformans*, *C. gattii* species complex) and lineages (*C. neoformans* lineages VNI, VNII, VNBI, VNBII). Species and lineage assignments were made based on quantifying the number of reads aligning to each reference genome within the concatenated reference file; this approach also allowed us to identify hybrid strains that exhibited strong matches to two reference genomes. Lineage assignments indicate that each patient was infected by a distinct haploid lineage (VNIa, VNIb, VNII), hybrid diploid lineage (*C. neoformans*/*C. deneoformans*, VNII/VNBII) or a mix of lineages (Figure 1, Table S1).

Following lineage assignment, reads from each strain were realigned to a lineage-specific reference or, in the case of hybrids, to a lineage-specific concatenated hybrid reference genome. Average genome-wide depth of coverage across the isolates was ∼48*×* (Table S2).

Aneuploidy and large duplications and deletions were inferred from normalized read depth estimates. Single nucleotide variants and small indels were called relative to the appropriate reference genome based on a haploid variant calling pipeline (Garrison & Marth, 2012; Li & Durbin, 2010). Detailed descriptions of the genome analysis pipelines are provided in Methods.

### 3.4 Chronic infections are characterized by complex genome restructuring

Four of the patients had chronic infections lasting between one and two and a half years. For these persistent infections, all but one of the strains isolated over the course of infection can be shown to be clonally related (within the patient) to the earliest isolated strain. Patient NC1 experienced a persistent infection lasting more than two years, caused by a lineage of VNIb strains that disseminated from the lungs (sputum) to the brain (CSF); NC8 was infected for over one year with a *A. neoformans*/*C. deneoformans* hybrid lineage; NC94 experienced a persistent infection for more than two years with a *C. neoformans* VNII/VNBII hybrid lineage which was first detected in their blood and subsequently in both the blood and brain (CSF); NC183 had a persistent pulmonary infection for over one year with a VNIa lineage which was first detected in a sputum specimen and later in a pulmonary nodule biopsy (Table S1). Except for patient NC183, evolution of the cryptococcal lineages within each of these patients was characterized by complex changes in genome architecture (aneuploidy, large chromosomal duplications and deletions, loss-of-heterozygosity, etc.) coupled with relatively modest accumulation of small-scale genetic variants (SNPs, small insertions/deletions).

### 3.5 Genomic diversification and branching evolution within a single patient

Samples from patient NC1 provided an opportunity to study chronic cryptococcal infection at great depth over a period of more than two years of infection. Specimens representing 13 collection time points over the course of infection were available from this patient, starting at 20 days of infection (DOI) and extending to DOI 855 (Table S1). Two or more phenotypically distinct strains were isolated from every specimen except at DOI 201. In total, 31 strains were isolated and sequenced from this patient. Analysis of the genome sequences of the NC1-derived strains indicates that they fall within the VNIb lineage and are very closely related to the *C. neoformans* reference strain H99 which was isolated at the Duke University Hospital in the 1978 (Janbon et al., 2014).

In order to better understand whether the isolates from NC1 were clonally related or representative of mixed infections (Desnos-Ollivier et al., 2010), we used allelic differences at single-nucleotide variant sites to estimate a phylogeny. Since we determined that the NC1 isolates were very closely related to H99, we included in our phylogenetic analyses H99 itself plus seven additional strains identified as most closely related to H99 in the study by Desjardins et al. (2017). A maximum-likelihood tree (RAxML-NG v1.2.2; (Kozlov et al., 2019)) of the NC1 isolates plus additional strains is shown in Supplementary Figure S7.

In the estimated phylogeny, all but one of the NC1-derived strains fall into a single very well supported sub-clade (100% bootstrap support) with very short branches (a near polytomy), consistent with the hypothesis that these strains are clonally related and likely derived from a single infectious strain. The exception to this is the strain MCM528-E, isolated at DOI 290, which is the strain most closely related to H99 among the strains analyzed (Figure S7). MCM528-E and H99 are nested within a well-supported clade of non-NC1 strains. MCM528-E was isolated from the same patient specimen as MCM528-F, one of the clonally related strains discussed previously. Our analysis thus provides strong evidence that patient NC1 had multiple independently derived infective strains for at least a short-period during their infection.

#### 3.5.1 Chromosomal sub-lineages in patient NC1

Based on read depth analysis of mapped genomic reads (Figure S18), the earliest NC1-derived strains isolated at DOI 20 have a standard *C. neoformans* haploid complement of 14 nuclear chromosomes. Though all strains except MCM528-E are clonally related, genome diversification within patient NC1 is characterized by extensive genomic restructuring and branching evolution and the establishment of genetically and phenotypically distinct sub-lineages. As described below, we discovered three major sub-lineages and several minor sub-lineages within patient NC1 over the course of ∼2.5 years of chronic infection. Each major sub-lineages persisted for an extended period of time and multiple sub-lineages were found within single patient specimens (Figure 3).

**Figure 3:**
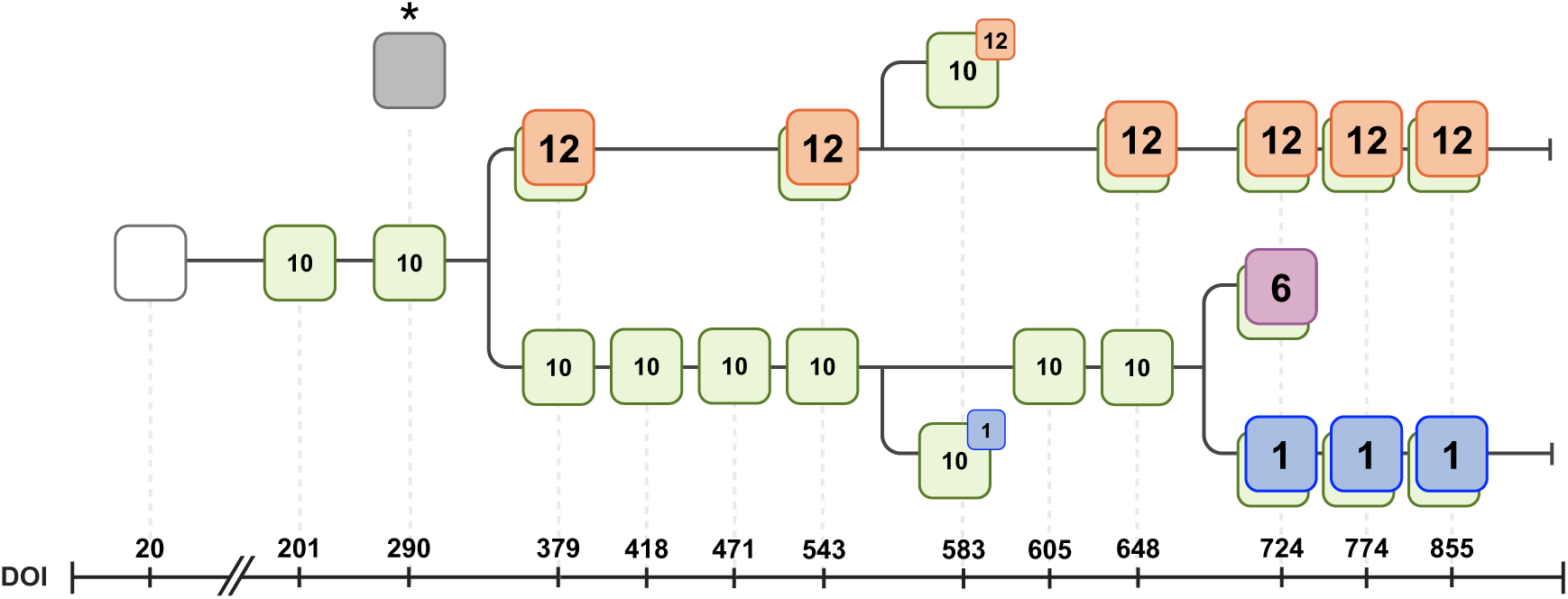
Chromosomal sub-lineages detected in patient NC1. Timeline of genomic changes in NC1 isolates during the course of infection. The white square represents the ancestral euploid (haploid) chromosomal arrangement in the earliest isolate. Green squares represent euploid genomes with an ∼11 kb deletion on chromosome 10. The dark grey square marked with an asterisk represents the single occurence of a euploid isolate (MCM528-E) that does not appear to be clonally related to the other isolates. Blue, orange and magenta squares represent genomes with chromosome 1, 12, and 6 aneuploidies, respectively. The branches at DOI 583 represent strains harboring a partial chromosome 10 deletion along with partial duplications of either chromosome 1 or 12. Each of the aneuploid lineages shares the deletion on chromosome 10.

The earliest structural genomic change in the NC1 lineage is a ∼11 kb deletion on chromosome 10, first detected at DOI 201. This deletion is predicted to result in partial truncation or complete loss of five genes, three of which are predicted to encode membrane bound sugar transporters proteins (Figure 4). The chromosome 10 deletion becomes fixed in the yeast population sometime between DOI 290 and 379; from DOI 379 on all strains isolated from NC1 share this deletion allele. The fixation of the chromosome 10 deletion is consistent with the hypothesis that loss of this genomic region is adaptive in this patient’s host niche, though we cannot rule out fixation due to hitchhiking with linked variants or neutral processes.

**Figure 4:**
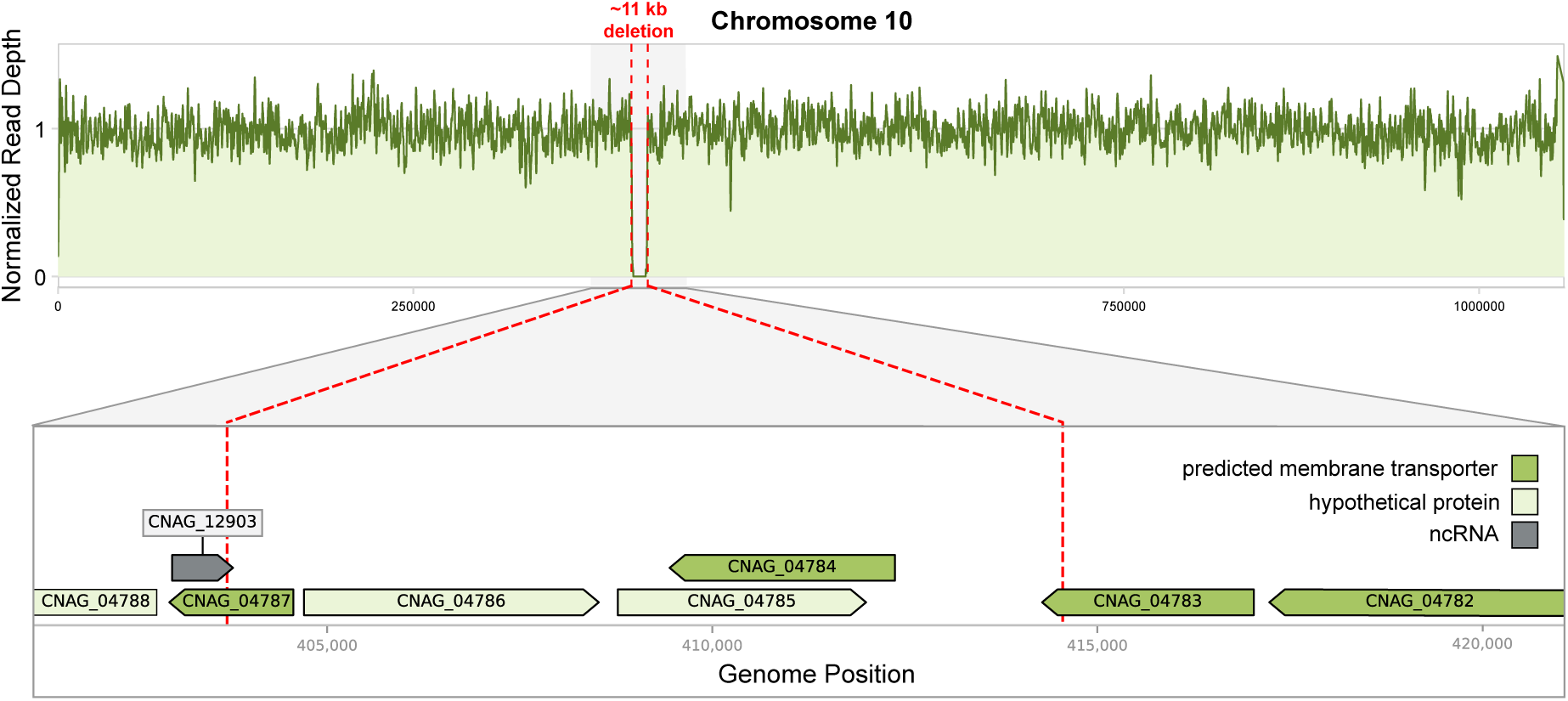
A deletion on chromosome 10 in strains of patient NC1. An ∼11 kb deletion of a region of chromosome 10 was first detected at DOI 201, located between positions 403,700 – 414,500 on the H99 reference genome. From DOI 379 on, all NC1 isolates share this deletion. Genes located partially or completely within this region include two hypothetical proteins and three predicted membrane transporters. Nearby genes include another predicted membrane transporter and a non-coding RNA sequence.

The second major genome change, first detected at DOI 379, is a duplication of chromosome 12 resulting in an aneuploid sub-lineage. Strains with chromosome 12 duplications persist until the last sample collection point (DOI 855). Although chromosome 12 aneuploidy was not detected at every time point between DOIs 379 and 855, likely due to limited sampling, we infer that this lineage was present throughout this period of the infection. At DOI 583, we detect a novel strain genotype with a partial chromosome 12 duplication. We hypothesize that this smaller duplicated region of chromosome 12 arose via partial loss of the whole chromosome 12 duplication. This partial chromosome 12 duplication genotype was not detected at any other time point.

The third major genomic change, a nearly complete duplication of chromosome 1, was first detected at DOI 724 and persisted until DOI 855. At DOI 855 a different chromosome 1 genotype is detected, in which a portion of the right arm has reverted to a single copy (Figure S19). This DOI 855 genotype appears to be derived from the earlier chromosome 1 lineage. As discussed below, we hypothesize that these sub-lineages with chromosome 1 duplications are selected for in response to inadequate antifungal therapy.

In addition to the three major chromosomal lineages described above, three other notable chromosomal genotypes appeared over the course of infection in NC1. At DOI 583 we recovered two different strains with partial duplications of chromosome 12 (∼190 kb duplication) and chromosome 1 (∼100kb duplication) respectively, and at DOI 724 a strain with a complete duplication of chromosome 6 was isolated. None of these three “minor” chromosomal sub-lineages was detected at any other time point.

DOI 648 is the last time point at which a euploid haploid strain was isolated from NC1. Every strain isolated from a subsequent time point possesses at least one aneuploid chromosome. The chromosome 12 sub-lineage thus co-existed for nearly 300 days with euploid strains. Following extinction of the euploid lineage, the chromosome 12 and chromosome 1 sub-lineages co-exist for 130 days.

#### 3.5.3 High impact mutations in NC1 isolates

We analyzed the genomes of the NC1-derived strains for genetic changes predicted to be “high-impact” with respect to gene function based on the variant annotation tool SNPEff Cingolani et al., 2012. Here we focus on a subset of the predicted high-impact variants that are shared across multiple strains in the NC1 lineage.

As described above, the 11 kb deletion on chromosome 10 that appears at DOI 201 and is fixed in the population by DOI 397 is predicted to lead to the complete or partial deletion of 5 predicted genes: CNAG_04783 – CNAG_04787 (Figure 4). Of these genes, CNAG_04783 and CNAG_04784 are predicted monosaccharide transporters (FungiDB R68; Alvarez-Jarreta et al., 2024). CNAG_04787 has no *Cryptococcus*-specific annotation, however the putative orthologs of this gene in other fungi are predicted to be ammonium permeases or acetate transporters (Paiva et al., 2004; Palková et al., 2002).

Three predicted high impact variants shared by all NC1 isolates, relative to the reference strain H99, are found in two sugar transporters (CNAG_05324, T482A; CNAG_06934, A149V) and an iron permease (FTR1 [CNAG_06242], I98L). These variants are also shared with the closely related non-NC1 derived strain A1-35-8, indicating that they arose prior to infection of NC1. CNAG_05324 and CNAG_06934 were previously found to be under selection in two population genomic studies (Desjardins et al., 2017; Sephton-Clark et al., 2022). Variants in FTR1 were significantly associated with fungal CSF burden in a study by Sephton-Clark et al. (2022).

Other notable genes with high-impact mutations in NC1 isolates include CDC1(CNAG_06647), YOX1 (CNAG_03229), SSK2 (CNAG_05063), and LMP1 (CNAG_06765). CDC1 encodes a phosphatase that Jin et al. (2020) identified as markedly upregulated during infection; we documented two different frame shift mutations in different subsets of the NC1 isolates. YOX101 is a homeobox transcriptional repressor that plays a role in oxidative stress response (Jung et al., 2015) and was found to be under positive selection in the population genomic analysis of Rhodes, Desjardins, et al. (2017). One NC1 isolate had a frameshift in SSK2 (CNAG_05063), which encodes a protein kinase involved in the HOG signaling pathway (Bahn et al., 2005). Prior molecular genetic and QTL mapping studies have shown that variation in SSK2 is associated with traits such as high-temperature growth and amphotericin B sensitivity (Bahn et al., 2007; Roth et al., 2021).

LMP1 is a particularly interesting gene, as it was first identified in the context of a study examining mutations that accumulated during laboratory passage of the reference strain H99 (Arras et al., 2017; Janbon et al., 2014). Janbon et al. (2014) found that *lmp1*Δ deletion mutants show reduced mating and melanization and are avirulent in mouse models of infection. The H99-derived laboratory with LMP1 loss-of-function mutations have been referred to as the “wimp” lineage (H99W) because of it’s highly attenuated virulence relative to wild-type H99 (Arras et al., 2017). It is therefore quite remarkable that we identified a frame-shift mutation (A335L) in two NC1-isolates, MCM300D and MCM300L, at the very last sampling timepoint (DOI 855). This frame-shift mutation leads to a premature stop codon at residue 344, truncating approximately 75% of the length of the predicted wild-type protein.

### 3.6 Hybrid lineages undergo extreme genome remodeling during chronic infection

Two patients, NC8 and NC94, were infected by diploid hybrid strains. NC8’s hybrid lineage was an inter-species hybrid, with chromosomal contributions from both *C. neoformans* and *C. deneoformans* ancestors. The isolates from NC94 were hybrids of the *C. neoformans* lineages VNII and VNBII.

Patient NC8 experienced a cryptococcal infection lasting 392 days prior to their death. Strains isolated from NC8 at DOI 0 have an approximately diploid chromosomal composition with equal contributions from both ancestral genomes. Exceptions to this are chromosome 10, for which two *C. deneoformans* chromosomes were detected with no contribution from *C. neoformans*, and chromosome 12, which is triploid, with two copies of *C. neoformans* origin and one copy of *C. deneoformans* origin. Additionally, there are a smattering of loss-of-heterozygosity (LOH) regions. From DOI 256 to DOI 392 we detect strains with large scale duplications and LOH on chromosomes 3, 6, 7, 8, and 12.

Patient NC94’s infection lasted 735 days and was caused by a VNII/VNBII diploid hybrid lineage. Only a single strain is available at DOI 0 – this initial hybrid has chromosomal contributions from both parent lineages for every chromosome except chromosome 4, which has two chromosomes of VNBII origin. Since only a single strain was sequenced from DOI 0, we cannot rule out additional heterogeneity in the initial infection but each of the strains collected at DOI 735 can be clearly related to the initial infective strain. Thus, we consider diversification from this single source as the most parsimonious explanation. At DOI 735, four strains are available and every one of these has a distinct genomic makeup (Figure 5). MCM450A, which was isolated from a blood specimen, is diploid for the VNII derived chromosomes 1 and 8, is haploid for chromosome 3 with a single VNII derived copy, and is partially diploid for chromosome 6 with both copies from the VNBII ancestor. Large scale deletions and LOH regions are observed on MCM450A chromosome 12. MCM450N, also isolated from the blood, has two copies chromosome 2 derived from VNII (whole chromosome LOH); there is complex LOH and regional duplication on chromosome 6; and there is partial LOH and duplication of chromosome 8. MCM531A, obtained from a CSF specimen, is diploid for the VNBII derived copies of chromosomes 4 and 8 and possesses a large LOH region on chromosome 2. MCM531C, also isolated from the CSF, exhibits large LOH regions on the left arms of chromosomes 2 and 13. An interesting contrast emerges when comparing the differences between the strains collected from the blood versus those collected from the CSF. Blood derived strains (MCM450A and MCM450N) show changes in VNII derived chromosomes and complex LOH/duplication patterns especially on chromosomes 6 and 8 while CSF derived strains (MCM531A and MCM531C) show changes predominately in VNBII derived chromosomes and large LOH regions specifically on chromosomes 2 and 13.

**Figure 5:**
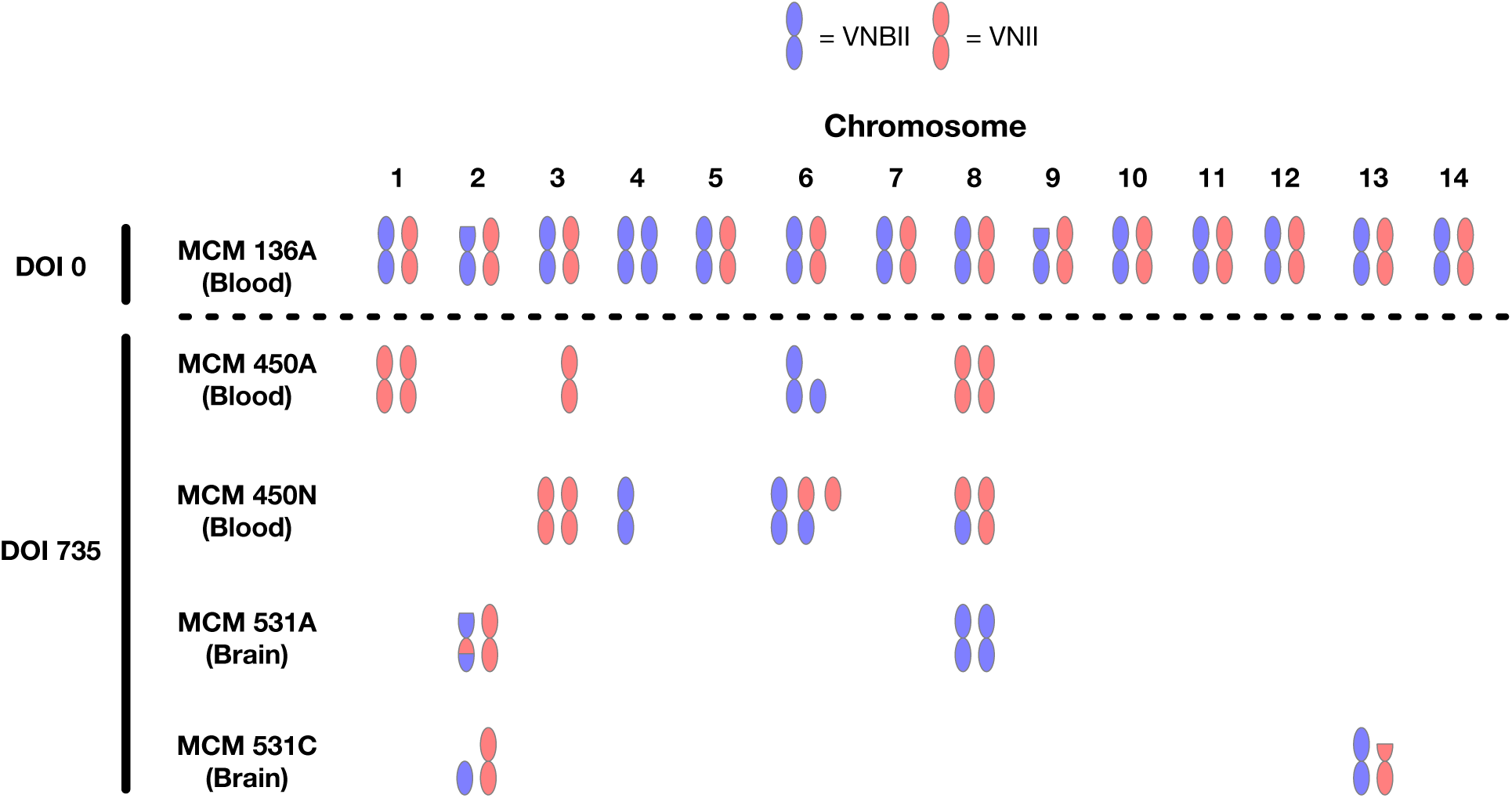
Structural variation in hybrid strains of patient NC94. Isolates from patient NC94 are available at DOI 0 and DOI 735. At DOI 735 every isolate sequenced has a different chromosomal structural arrangment. Variation includes chromosome loss, deletion of chromosome arms, and loss-of-heterozygosity.

### 3.7 Long-term recurrent infections are caused by reinfection

Cryptococcal infections can be recurrent, i.e. a patient who was previously thought to have resolved their infection may be re-diagnosed with a cryptococcal infection after some period of time (Garcia-Hermoso et al., 1999; Spitzer et al., 1993). In prior studies, recurrent infections presenting after 3-4 months of symptom free care were inferred to have been caused by relapse based on the observation that the strains from the recurrence were shown to be clonally related to the initial isolates (Spitzer et al., 1993). Cryptococcal persistence leading to relapse may be facilitated by cellular mechanisms that promote dormancy (Alanio, 2020; de Castro et al., 2023).

Two of the patients analyzed here allowed us to explore the question of relapse versus reinfection over very long time periods. The specimens from patient NC83 were collected almost 10 years apart; those from NC6 were separated by nearly 20 years. For both of these patients, genome sequences of strains from the initial and subsequent infections indicate that each patient’s second bout of cryptococcal disease was caused by reinfection with new strains. NC83’s initial infection involved a VNII strain, and their subsequent infection was caused by a VNIb strain. NC6’s incident infection was a VNIb strain with the subsequent infection caused by a VNIa strain. With respect to phenotypes such as colony texture and melanization, there are significant differences between strains from the early and late infections, suggesting there is no single combination of phenotypes that facilitates infection of a given patient. Importantly, both HIV-positive patients had completed their induction, consolidation, and maintenance antifungal therapy and had consistent antiretroviral therapy after the incident cryptococcal infection. For both patients, a lapse in anti-retroviral therapy following life events led to a window of increased susceptibility and a subsequent cryptococcal re-infection.

### 3.8 Phenotypic diversification accompanies genomic remodeling during chronic infection

*Cryptococcus* pathogenicity and virulence are influenced by traits such as growth rate, cell size and morphology, the thickness of the polysaccharide capsule, colony morphology, melanin formation and CO_2_-tolerance (Fernandes et al., 2018; Goldman et al., 1998; Iyengar & Xu, 2022; Jezewski et al., 2023; Rivera et al., 1998). For each of the chronic infection lineages we characterized these virulence-related traits and found that the extensive genomic evolution we observed over the course of chronic infection is accompanied by an explosion of phenotypic diversification and heterogeneity among the evolving sub-lineages. These trends are described below.

#### 3.8.1 Phenotypic evolution in patient NC1

Over time, strains isolated from patient NC1 show a general increase in the frequency of mucoid colony textures in response to nutrient limitation or high temperature and CO_2_ conditions (Figure 6A). These same strains showed a time-dependent increase in melanin formation rate with or without high CO_2_ (Figure 6B).

**Figure 6:**
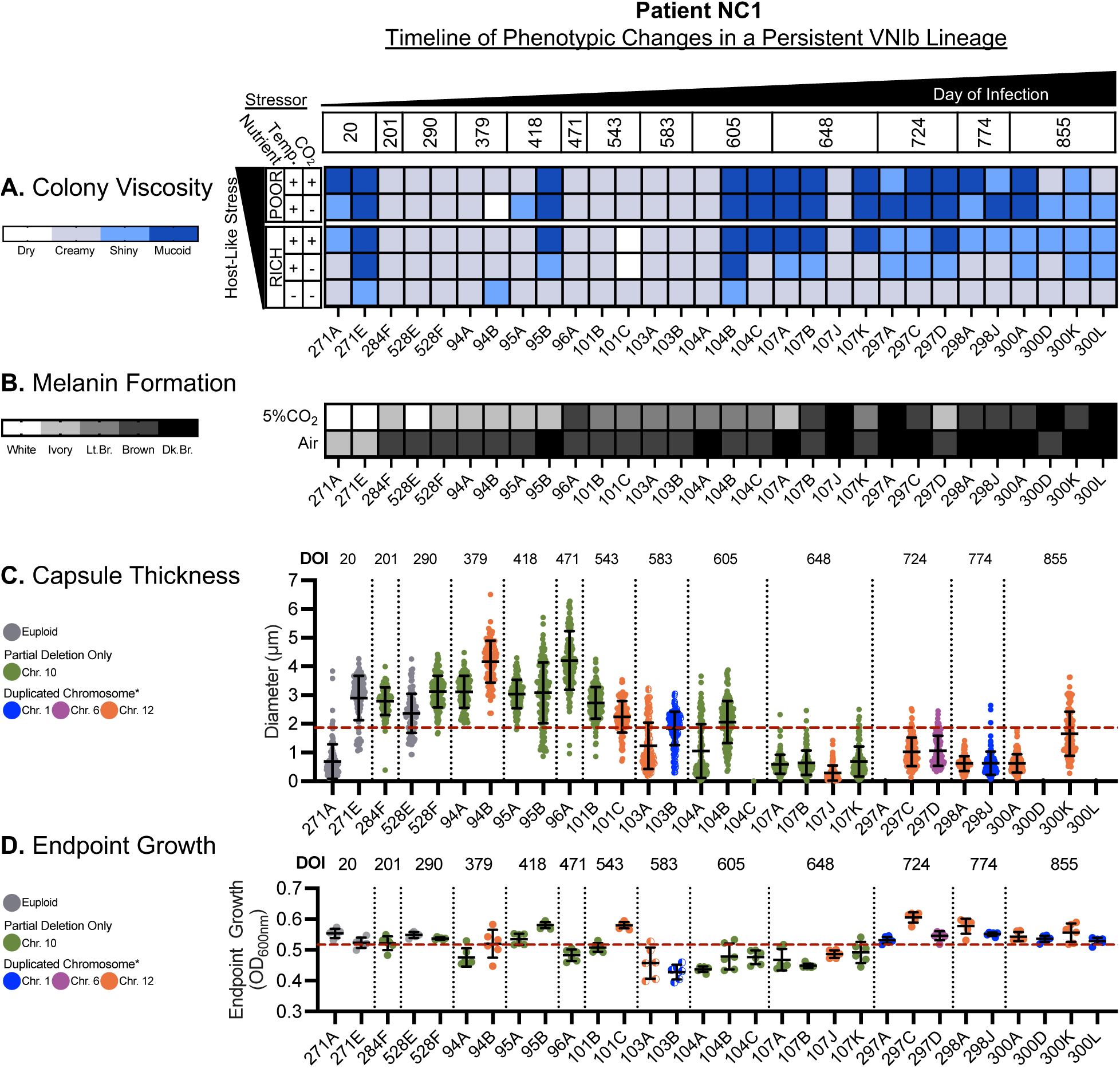
Phenotypic changes in patient NC1 isolates over time. The top x-axis depicts the timeline of persistence within the host, with columns corresponding to patient strains labeled on the bottom x-axis. A) Over time, the persistent NC1 lineage becomes increasingly mucoid under host-like stressors. B) When grown on melanin-inducing agar for 120 hours, strains within the persistent VNIb lineage that were collected during early infection exhibit a CO2-dependent delay in melanin formation that changes over time; becoming capable of producing melanin more rapidly in both ambient air and 5% CO_2_ conditions. C) When grown under host-like stress conditions (DMEM, 37°C, 5% CO_2_, 72h), the persistent VNIb strains from patient NC1 exhibit a time-dependent reduction in capsule thickness; where later strains have smaller capsules. Red horizontal dashed line represents the average capsule thickness of all measured strains (Mean = 1.67 um, SEM = 0.2075 um). D) Endpoint growth of persistent VNIb strains vary over time. Despite being smaller in size, later strains display similar endpoint growth compared to early strains suggesting an increase in growth rate and different to strains collected during the middle of NC1’s infection (DOI 583 – 648)

Capsule thickness of the NC1 strains showed a marked time-dependent decrease (Figure 6C), while cell body size shows little change (Figure S11). As a result, the total cell diameter (cell body size + capsule thickness) decreases over time. The decrease in capsule thickness is particularly notable starting at DOI 583. Several time points exhibit substantial heterogeneity in capsule size between sub-lineages isolated from the same patient specimen. In addition to the reduction in capsule thickness, NC1 sub-lineages with chromosomal duplications exhibited a high frequency of pseudohyphal or tear-drop shaped cell morphologies (Figure S10)

Endpoint growth rates in NC1 strains exhibit modest variability over time, but no strong trends (Figure 6D). Interestingly, strains with whole chromosome duplications of chromosomes 1, 6, and 12 do not manifest fitness defects relative to their euploid progenitor as is often the case in aneuploid genotypes. However, strains with partial chromosome duplications of chromosome 1 and 12, detected only at DOI 583, do exhibit decreased endpoint growth relative to the DOI 20 euploid genotype.

#### 3.8.2 Phenotypic evolution in hybrid lineages

The *C. neoformans*/*C. deneoformans* hybrid strains from patient NC8 exhibit significant variability in the thickness of the polysaccharide capsule, both within and between time points. However, unlike NC1, there is no trend towards thinner capsules over time (Figure S13 and S14). With respect to cell body size, NC8 strains are relatively similar both within and between time points. Endpoint growth in NC8 is variable among strains, but without any consistent temporal trends or obvious correlates with the chromosomal changes observed.

The VNII/VNBII hybrid strains in NC94 show relatively constant cell body size, but significant decreases in cell capsule thickness between DOI 0 and DOI 735 (Figure S15 and S16). The two strains isolated from a blood sample on DOI 735 have the smallest capsules, while the CSF isolates from the same time point have capsules somewhat closer in size to the original isolate. The two DOI 735 CSF isolates have modestly higher endpoint growth rates than the blood isolates than the three strains isolated from blood on DOI 0 and 735.

On the whole, the diploid hybrid lineages isolated from patients NC8 and NC94 exhibit significant phenotypic variation both within and between time points, but unlike the NC1 lineages they have less obvious trends.

### 3.9 Changes in antifungal susceptibility during in host chronic infections

The standard of care for cryptococcal disease is administration of amphotericin B (AmB), flucytosine (5FC), and/or the azole antifungals (most commonly fluconazole (FLC)) (Chang et al., 2024; Perfect et al., 2010). Each of the patients included in our analysis of chronic cryptococcosis received a combination of these drugs, though with variable days of exposure, dosages, and continuation of the prescribed regimen after discharge.

For each strain we measured growth inhibition by AmB and FLC in two ways: 1) classical minimum inhibitory concentration (MIC) determination by visual inspection and 2) measurement of endpoint growth by optical density (OD_600nm_). Optical density measures across multiple drug concentrations were combined to estimate dose-response curves for each drug and the area under the curve (AUC) for the dose-response curves was used as a metric of antifungal susceptibility. Below we describe the detailed relationship between fungal genotypes, antifungal treatment regimens, and susceptibility to antifungals for each strain.

#### 3.9.1 Antifungal susceptibility of patient NC1 isolates

Over the course of their ∼2.5 years of cryptococcal infection, patient NC1 received AmB, 5FC, and FLC. However, this patient experienced inadequate and sporadic antifungal therapy during hospital admissions and had challenges continuing antifungal therapy after discharge (Supplemental Case Report - NC1). Figure 7 depicts the estimated days of antifungal exposure over time for this patient based on documented instances of antifungal administration from their electronic medical records. After the initial encounter at which the patient received 5 days of AmB + 5FC treatment and 6 days of FLC therapy, there was no additional antifungal treatment until after day 200. After DOI 201 a more regular, but still inadequate, regimen of AmB + 5FC exposure began during CM-related admissions. Unfortunately, antifungal therapy was not provided during every CM-related admission. Furthermore, patient NC1 never completed the standard two-weeks of AmB+5FC induction therapy until their final CM-related admission. Patient NC1’s initial exposure to FLC was minimal and did not resume until after DOI 397, again inconsistently, and from DOI 543 to DOI 855 no additional FLC treatment was received. (Figure 7).

**Figure 7:**
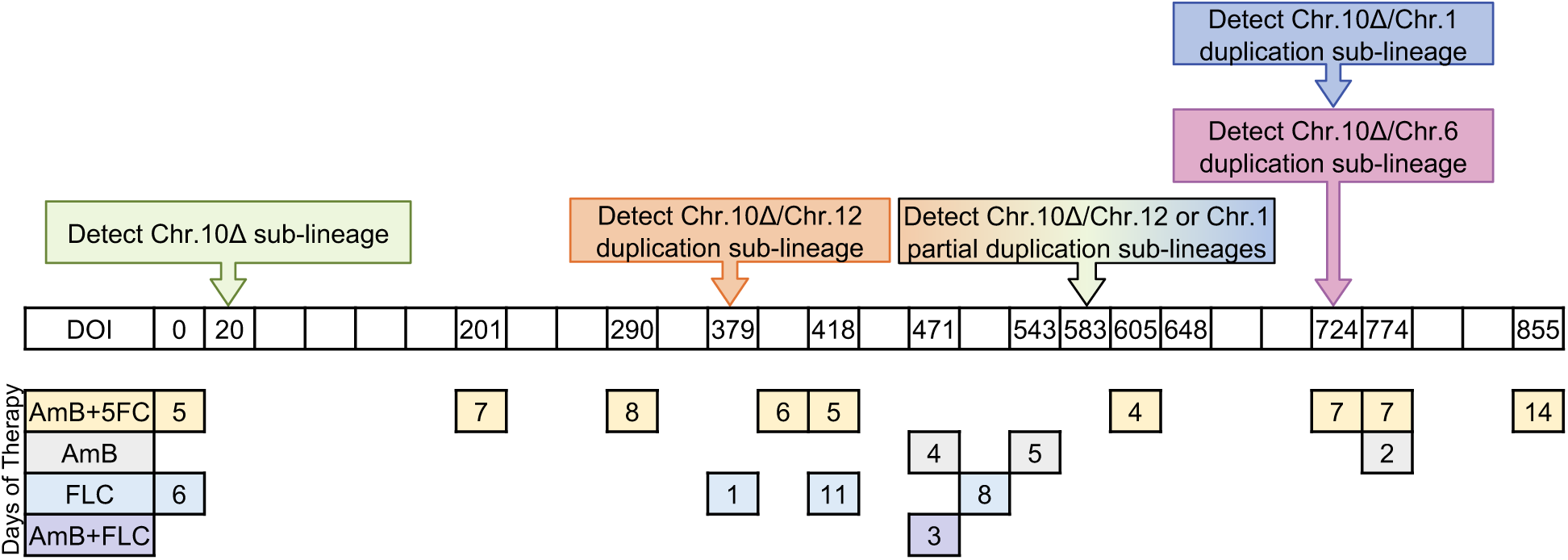
Cumulative days of antifungal drug exposure for patient NC1. Patient NC1 received AmB, 5FC, and FLC over the 2.5 years of their recurrent infection. This figure depicts the number of days of drug treatment received at various period over the period of infection.

Inadequate and sporadic antifungal therapy can select for mutations that increase resistance or reduce susceptibility. In *Cryptococcus* and many other fungal pathogens, chromosomal duplications and deletions have been frequently found to confer resistance to antifungal drugs (Selmecki et al., 2009; Sionov et al., 2010). We therefore compared antifungal drug responses between euploid and aneuploid strains isolated from NC1 (Figure 6).

NC1 isolates with the chromosome 10 partial deletion and those from the chromosome 12 aneuploid lineage exhibit a modest decrease in susceptibility to AmB compared to the earliest euploid strains (Figure 6A,C). Strains from the NC1 chromosome 1 sub-lineage show markedly reduced susceptibility to FLC compared to other strains isolates from NC1, consistent with prior studies implicating chromosome 1 duplications with azole tolerance (Figure 8). However, in contrast to prior work that found a fitness cost of chromosome 1 aneuploidy, we did *not* detect major growth defects for these strains under in vitro host-like growth conditions in the absence of FLC (Figure S10-S11).

**Figure 8:**
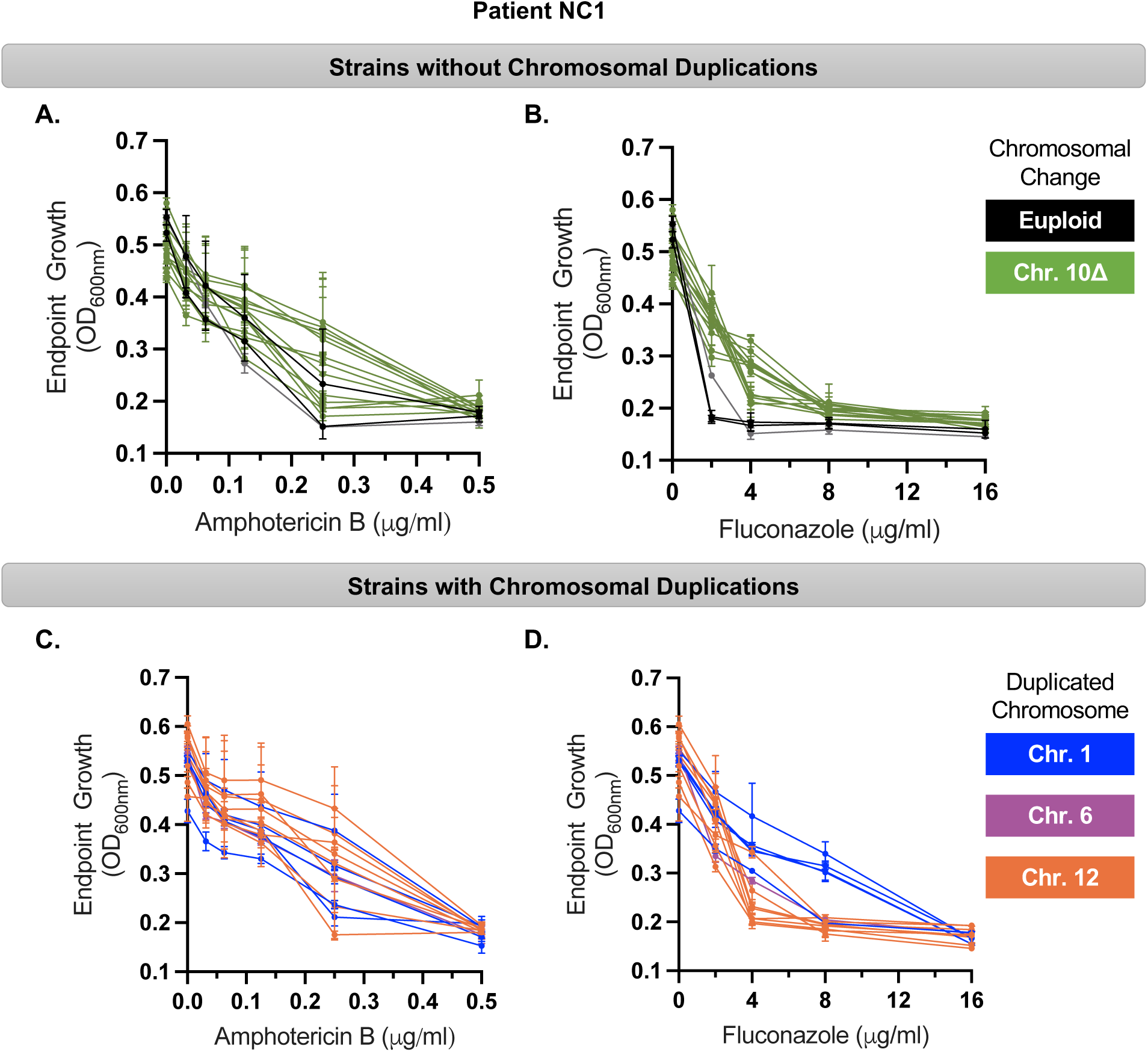
During persistent CNS infection antifungal susceptibility decreases with and without large chromosomal duplications. A, B) NC1 strains that do not contain chromosomal duplications, but harbor a partial deletion in chromosome 10, display a reduction in antifungal susceptibility to both Amphotericin B and Fluconazole. C,D) NC1 VNIb strains with chromosomal duplications display a slight reduction in antifungal susceptibility to both Amphotericin B and Fluconazole. However, the strains harboring a chromosomal 1 duplication have a substantial decrease in fluconazole susceptibility compared to all other strains.

#### 3.9.2 Antifungal Susceptibility of Hybrid Strains

Patient NC8’s persistent *C. neoformans*/*C. deneoformans* hybrid isolates showed no change in AmB MIC over the course of infection despite having the longest exposure to AmB (71 days) relative to all other patients. The FLC susceptibility, however, did show a significant change where earlier strains had an MIC of 16 µg/ml and later strains had an MIC that ranged from 32 µg/ml to 64 µg/ml. This change in FLC susceptibility was evident in the analysis of endpoint growth, where later strains exhibited greater growth at lower concentrations of FLC (Figure S14).

For patient NC94 strains, antifungal susceptibility to AmB showed little change in MIC (0.125 – 0.25 µg/ml) for strains collected between day 0 to day 735, perhaps due to limited AmB exposure. However, despite having extensive exposure to FLC (542 days), antifungal susceptibility to FLC also remained stable, with a MIC of ≤2 µg/ml (Table S1, Figure S16).

## 4 Discussion

The case studies we describe here represent a rare opportunity to dissect fungal evolution during chronic infections; though the number of cases is modest, they yield novel insights about fungal evolution in response to long-term occupancy of human hosts.

### 4.1 Chronic cryptococcal infections derive from diverse lineages

From the six patient cases analyzed, it is clear that chronic cryptococcosis can be caused by many different *C. neoformans* lineages. This patient cohort had little reported travel outside of the southeastern USA, however every one of the infections we analyzed was derived from a distinct lineage. Strains assigned to the VNI molecular type are the most frequently reported lineage in acute infections (Cogliati, 2013) but only two of the six patients we examined had infections caused solely by strains of VNI origin.

Two of the six patients had infections caused by hybrid strains – one an interspecific hybrid and the other an intraspecific hybrid. The intraspecific hybrid, a VNBII/VNII diploid strain, is particularly interesting as pure VNBII strains have never been reported from North America. In the Americas, hybrid strains with chromosomal contributions from the VNB lineages have been reported only in the case of interspecific hybrids (Litvintseva et al., 2007). Rhodes, Beale, et al. (2017) reported VNBII/VNII hybrids, but only from Africa. The origins of hybrid strains with VNB ancestry, and the contribution of the VNB lineage to *Cryptococcus* diversity in the Americas are thus of significant interest.

### 4.2 Chronic infections are characterized by extensive genome restructuring

The most striking changes we observed in strains isolated from patients with chronic infections was a high degree of genomic structural variation, including whole chromosome duplications, large and small deletions and duplications, loss-of-heterozygosity, and likely rearrangements. In only one of the patients, NC183, do we find no evidence of genome restructuring. Interestingly, this patient differed from the others in being a solid organ transplant patient, rather than a person with HIV/AIDS. Viral evolution has been previously shown to occur at a high rate in persons with immunocompromising conditions (Ko et al., 2024); as more data become available it will be interesting to see if this is true with respect to fungal and other microbial infections.

Aneuploidy has been shown to be an important mechanism of genomic variation in diverse fungi (Bennett et al., 2014) and likely contributes to adaptation to antifungal drugs (Selmecki et al., 2009). Chromosomal duplications are found with a relatively high frequency among *C. neoformans* clinical isolates (Chen et al., 2017b; Hu et al., 2011; Rhodes, Beale, et al., 2017; Sionov et al., 2010). For example, chromosome 12 is among the most commonly duplicated chromosomes in VNI strains and such duplications have been previously recovered from serial isolates(Chen et al., 2017b; Ormerod et al., 2013; Rhodes, Beale, et al., 2017; Sephton-Clark et al., 2022). No specific phenotypes have been definitively tied to duplication of chromosome 12, though Rhodes, Beale, et al. (2017) suggest that these duplications are a response to fluconazole stress. We detected no significant difference in fluconazole susceptibility in NC1 strains with chromosome 12 aneuploidy, but such strains are among the least susceptible to amphotericin B, which is the other major antifungal prescribed to most patients with cryptococcal disease. Notably, a recent study from our laboratory detected a large effect amphotericin B QTL on chromosome 12 in a genetic cross of *C. deneoformans* strains (Roth et al., 2021). This chromosome 12 QTL also had pleiotropic effects on high-temperature growth.

*C. neoformans* duplications of chromosome 1 are associated with selection under azole stress, presumably due to dosage effects of key genes such as ERG11 and AFR1 that are important for azole resistance and that are found on chromosome 1 (Sionov et al., 2010). Our findings from patient NC1 are consistent with these prior studies – the sub-lineage with chromosome 1 duplications showed a large decrease in susceptibility to fluconazole compared to other lineages. Chromosome 1 aneuploidy is usually considered unstable, and several prior studies have documented a high rate of reversion to euploid copy number when selection is relaxed (Sionov et al., 2010). It is notable that the chromosome 1 aneuploid sub-lineage was not detected until quite late in patient NC1’s course of infection, being first detected at DOI 724. DOI 724 is nearly 200 days after FLC treatment had stopped and the chromosome 1 duplicated lineage continued to persist until the patient died on DOI 885. The chromosomal 1 aneuploids we found were also stable under laboratory passage in rich media, and we detected no growth defect relative to the euploid ancestor when comparing growth in stressful conditions meant to mimic aspects of the host environment. This suggests the possibility that there may be compensatory mutations that ameliorate the deleterious effect of chromosome 1 aneuploidies.

The two diploid hybrid lineages, found in patients NC8 and NC94, provide insight into the prevalence of large scale loss-of-heterozygosity. Loss-of-heterozygosity has been frequently observed in pathogenic fungi, such as *Candida albicans*, that are primarily diploid (Zhou et al., 2024) as well as non-pathogenic species such as *Saccharomyces cerevisiae* (Magwene et al., 2011). *C. neoformans* is considered a “preferentially haploid” organism but both hybrid and non-hybrid diploid Lin et al., 2009 strains have been detected at appreciable frequency (5-30%) in some studies. In diploid species that are asexual, or experience rare meioses, mitotic recombination induced LOH has been hypothesized to be an important contributor to standing variation and adaptation (Mandegar & Otto, 2007; Smukowski Heil, 2023). The high frequency of LOH in the two hybrid lineages described here may have contributed to the adaptation of these lineages within their host niches.

### 4.3 An explosion of phenotypic heterogeneity accompanies genomic diversification

Similar to the genomic changes we observed, chronic infections were also characterized by a high degree of phenotypic heterogeneity both between and within serial samples. Some of the temporal phenotypic trends we observed, such as decreased antifungal susceptibility and increased melanization, were expected. However, other phenotypic changes, such as a decrease in capsule thickness, were surprising given current understanding of such traits in promoting virulence in animal models. It is important to note across the six patients analyzed we detect no global trends, consistent with the idea that fungal adaptation during pathogenesis may be both complex (see below) and highly host specific.

### 4.4 Branching evolution and co-persistence of infective lineages

Our study demonstrates that during chronic infection, multiple genotypically and phenotypically distinct sub-lineages arise and co-persist over extended periods of time (months to years). For example, in patient NC1, a sub-lineage characterized by chromosome 12 aneuploidy branched off a euploid lineage, and then co-existed with that euploid and other independently evolve aneuploid sublineages for at least 475 days. Not only do sub-lineages overlap, but they are found in the same patient specimens suggesting that these lineages occupy the same tissues or organs.

The pattern of multiple genotypically and phenotypically distinct lineages and co-existing over-time is consistent with models of niche specialization and adaptive radiation in heterogenous environments (Rainey & Travisano, 1998) and/or clonal interference (Gerrish & Lenski, 1998; Kao & Sherlock, 2008; Lang et al., 2013; Selmecki et al., 2015). We hypothesize that the complex host-environment may not select for single “most fit” genotypes; rather there are likely multiple host sub-niches where fitness is dependent on different combinations of traits. It is also possible that interactions between sub-lineages may promote coexistence through mechanisms such as metabolic cross-feeding (Wang et al., 2023) or collective resistance/tolerance to antifungals (Bottery et al., 2021).

Our findings are consistent with other recent studies that have documented co-persistence of distinct subpopulations during long-term fungal infections. For example, Demers et al. (2018) documented distinct sub-populations of *Candida lusitaniae*, derived from a recent common ancestor, in different lobes of the lungs of a cystic fibrosis patient. The distinct populations showed both genotypic and phenotypic heterogeneity, including differences in drug-resistance. Genomic changes in these *C. lusistaniae* sub-populations included both small scale and large scale changes such as aneuploidy of several chromosomes. Similarly, Ross et al. (2021) documented the co-existence of both transitory and persistent lineages of *Aspergillus fumigatus* in a 4.5 year time series of a single cystic fibrosis patient.

### 4.5 Socioeconomic factors contribute to the development of chronic cryptococ-cosis

Risk of infection, as well as the effectiveness of antifungal treatment, relies heavily on a patients’ access to equitable healthcare and medication. Socioeconomic factors, known as social determinants of health (SDOH), influence health outcomes and are broadly categorized into eight key domains: 1) Socioeconomic Status; 2) Education; 3) Employment and Working Conditions; 4) Neighborhood and Physical Environment; 5) Social Support and Networks; 6) Healthcare Access and Quality; 7) Cultural Factors; 8) Discrimination and Social Justice (Braveman & Gottlieb, 2014; Islam, 2019). These domains are interconnected and real-world issues often span multiple categories. Collectively, they shape patients’ health trajectories, directly impacting their ability to access consistent and comprehensive care.

Among the six chronic cryptococcosis patients, all five male patients exhibited high-risk SDOH that may have contributed to their chronic and persistent cryptococcal infections. The most frequently documented risk factors included untreated mental health conditions, substance abuse, homelessness, lack of support systems, history of incarceration, access to healthcare, and health-care worker bias. These factors are closely linked to challenges in disease prevention, detection, clinical care, adequate antifungal therapy, and health outcomes. Focusing on challenges in continuing antifungal therapy (AFT) after discharge, patients NC1, NC8, and NC94 experienced housing instability and/or homelessness during their chronic infections. However, when these patients had support systems—such as home health services, a family/friend caregiver, or access to a homeless shelter that provided medications, including antifungals—they were more likely to adhere to AFT and anti-retroviral therapy (ART), attend regular ID clinic appointments, and experienced longer intervals between severe CM symptoms and re-hospitalization.

### 4.6 Epidemiological challenges for identifying chronic cryptococcosis

Epidemiological studies of cryptococcosis, outside the context of clinical trials, typically rely on diagnostic codes within electronic medical records to identify cases. However, this approach is susceptible to inconsistencies in coding practices among healthcare professionals, leading to missed cases, as observed in our study of the six chronic cryptococcosis patients. For example, patient NC183 had *Cryptococcus*-positive sputum and pulmonary nodule cultures but lacked any formal *Cryptococcus*-related diagnoses in their medical records. In patient NC6, only 3 out of 6 health care encounters had a *Cryptococcus*-related diagnosis, including two encounters in which the patient was culture-positive and one without a positive cryptococcal diagnostic test. Patient NC83 had 3 out of 18 hospital admissions with *Cryptococcus*-related discharge diagnoses: three primary CM (two encounters in which the patient was culture-positive and one encounter where the patient was culture-negative but their CSF CrAg titer was 1:512). Finally, patients NC1, NC8, and NC94 consistently had a primary diagnosis of CM during encounters and they were culture-positive for *Cryptococcus*. Our analysis of these cases reveals variations in coding practices across time and among different clinicians.

### 4.7 Implications for clinical care and diagnosis of chronic fungal infections

Both our analysis of chronic cryptococcal disease, as well as several other recent investigation of prolonged fungal disease (Demers et al., 2018; Murante & Hogan, 2024; Ross et al., 2021), demonstrate that such infectious often involve genetically and phenotypically diverse sub-populations that evolve and co-exist over long time periods. Clinically relevant sub-populations, such as drug-resistant genotypes, may be stable throughout an infection even if their frequency is low (Demers et al., 2018). Unfortunately, clinical diagnostic practices typically end at cryptococcal species identification. In the rare instances where antifungal susceptibility testing is performed, the standardized protocol uses a single colony rather than a population thereby limiting our understanding of fungal susceptibility in patients with multi-strain infections. Additionally, the recognition that phenotypic heterogeneity can exist within a genotypically homogeneous population often leads to biased genotype isolation from clinical samples. These practices limit the detection of sub-lineages or mixed lineage infections, both in the clinic and in research using clinical isolates. As a result, our understanding of the pathogenic advantages and clinical implications of genotypically and phenotypically diverse infections remains limited.

Our analysis of clinical data from these chronic cryptococcosis cases highlights real-world factors that may increase the risk for chronic infection, beyond the yeast’s ability to adapt. Notably, our findings from serial specimens collected over 20 years from patient NC6 and over 10 years from NC83 underscore the essential role of anti-retroviral therapy in preventing recurrent cryptococcal infections in patients with HIV/AIDS. This case series also reflects many previously reported risk factors associated with chronic cryptococcosis, such as inadequate antifungal induction and maintenance therapy, ineffective fungal clearance during initial treatment, prolonged immunosuppression from illness or medication, and continued environmental exposure due to socioeconomic factors like homelessness. Specifically, in patient NC1’s case, inadequate antifungal therapy may have provided symptom relief, but was not sufficient to eradicate the infection. Access to equitable clinical care, ensuring proper ART and AFT, is thus crucial for preventing reinfection and achieving a complete cure after initial diagnosis.

Although our sample size is limited, we hypothesize that equitable care—which improves access to medications and support for ART and AFT adherence outside of the hospital—has a positive impact on outcomes in chronic cryptococcosis, similar to other infectious and chronic diseases in individuals with high-risk SDOH. Our findings, particularly the long-term data from patients NC6 and NC83, emphasize the critical role of consistent ART in preventing recurrent cryptococcal infections and enabling early detection and treatment of opportunistic diseases. While specific correlations between SDOH and health outcomes cannot be definitively established in this small cohort, it remains essential to consider SDOH as significant factors affecting a patient’s ability to complete AFT and prevent cryptococcosis relapses.

An increased appreciation of the potential complexity of fungal infections is critical for effective diagnosis and treatment of chronic fungal disease, and it is our hope that this and future studies will lead to changes in clinical diagnostic practice and encourage physicians to adopt a SDOH-focused approach to equitable care practices that will both increase the probability of detecting such cases, prevent subsequent infections, and improve health outcomes.

### 4.8 Ecological and evolutionary dynamics in heterogeneous infections: The next frontier

The long-term persistence of genotypically and phenotypically distinct *Cryptococcus* lineages within single patients begs the question of how such lineages manage to co-exist. Do such lineages occupy spatially distinct regions of host tissues or organs? Is there metabolic differentiation that allows such lineages to utilize different host resources? Are interactions between lineages relevant to their fitness, both individually and as a group? How do such interactions change dynamically, over time or as a function of population density or host status? Questions such as these, and how they relate to in-host evolution of pathogen populations, are fundamentally questions of evolutionary ecology. We look forward to the wealth of novel insights and new discoveries to come as medical mycology and microbial evolutionary ecology become increasingly intertwined in pursuit of answers to these important questions.

## 5 Online Methods

### 5.1 Study population and clinical cohort

This case series report represents a retrospective examination of Duke University Health System (DUHS) patients that had a *Cryptococcus*-positive culture collected between January 1, 1991 – December 31, 2022. The specific chronic cryptococcosis patient cohort was identified from the Duke University clinical lab mycology isolate collection. Inclusion criteria were: 1) patient must have at least two available culture-positive clinical specimens collected at two different timepoints at least 365 days apart; 2) patient must have electronic medical record clinical information available; and 3) patient must be at least 18 years of age at the time of specimen collection. Clinical information for each patient was collected for each clinical encounter when the patient tested positive for *Cryptococcus*. Clinical data were recorded by performing chart reviews of electronic medical records through the use of a standardized and comprehensive case report form (file in supplement). Clinical information that was collected included demographics, social history, past medical history, and clinical course of the patient (symptoms, diagnostic tests, medications, and outcomes) throughout the duration of serial cryptococcal infections (Perfect et al., 2010). Patient charts were comprehensively reviewed from first healthcare encounter until death or up to 10 years after their last *Cryptococcus*-positive diagnostic test. In addition to a physical case report form, the clinical information was stored digitally in a REDCap database and analyzed using SAS statistical software.

Clinical isolates of *Cryptococcus*, along with associated clinical information and generated data, were obtained in compliance with ethical guidelines and ethical approval was given by the Duke University Medical Center Institutional Review Board (Durham, North Carolina) for the following protocols: Pro00005314 (Database and Specimen Repository for Infectious Disease-Related Studies), Pro00001304 (Treatment of Cryptococcosis in the Era of Lipid Amphotericin B Formulations), and Pro00020345 (Treatment of Cryptococcosis in the Era of Lipid Amphotericin B Formulations: data from January 1, 1930-December 31, 2024).

### 5.2 Yeast strains and General Microbiological Methods

The specimens collected from the six patients with chronic cryptococcosis in this study were stored by clinical laboratory technicians in the process of routine clinical care without adherence to a standardized procedure. Consequently, the stored specimen stocks for each patient could encompass a variety of organisms (bacteria, yeast, filamentous fungi) and/or a variety of genotypically distinct cryptococcal colonies cultured directly from the patient without purification is more common. In some cases, specimens might have undergone purification to isolate *Cryptococcus* as either a single colony or multiple colonies (less common). In instances where laboratory technicians undertook purification prior to storage, there may have been a potential bias in colony selection, favoring those adhering to the classical *Cryptococcus* colony phenotype which would result in an underestimation of strain diversity.

All fungal strain glycerol stocks were stored at -80°C until time of use. Prior to experiments, strains were propagated at 30°C for 48 - 72 hours on yeast peptone dextrose (YPD) agar plates. Experiments were performed within 10 days of propagation.

The unpurified *Cryptococcus* clinical specimens were obtained from the Duke University Hospital Clinical Laboratory Fungal Archive Collection. These unpurified clinical specimens were streaked for individual colonies on YPD agar and the melanin-inducing agar, L-DOPA and incubated at 30°C or 37°C for 72 hours, respectively. Patient specimens that exhibited phenotypic heterogeneity among the yeast colonies when grown on YPD and/or L-DOPA solid media underwent phenotypic validation by patching at least three colonies from each distinct phenotype onto YPD and L-DOPA agar. Phenotypically distinct colonies that displayed stable phenotypes were saved for further analysis in addition to the original mixed specimen. During specimen screening, multiple genotypically distinct colonies were isolated from the unpurified specimen stock, labeled with the original strain ID (e.g. MCM125) followed by a letter to distinguish individual colonies (i.e., MCM125A, MCM125B, MCM125C). Clinical specimens that grew *Cryptococcus* and bacteria or other non-*Cryptococcus* fungi were purified by picking and patching at least ten yeast colonies and verifying the cryptococcal identity via melanin formation and induction of polysaccharide capsule. Patient specimens that did not display phenotypic heterogeneity among yeast colonies on YPD and L-DOPA agar underwent phenotypic verification of cryptococcal identity and were stocked as a homogeneous population. All clinical specimens were validated to be *Cryptococcus* by melanin formation and India ink stain. Unless otherwise indicated, experiments began with an initial culture of purified yeast cells that were grown overnight in YPD liquid media, shaking at 30°C.

### 5.3 In vitro Antifungal Susceptibility and Cell Growth Assays

All antifungal drugs were solubilized in DMSO. Antifungal susceptibility testing was performed using the protocols described in the document CLSI M27-A3 with a modified temperature of 37°C to better replicate the human host environment and direct counting of yeast innocula (CLSI, 2008). The reported minimum inhibitory concentration (MIC) values represent the highest value for at least three independent biological replicates performed in technical duplicate. MIC results were read by visual inspection and by cell optical density readings at absorbance 600 nm (OD_600_); yeast cells were resuspended before absorbance reading. Endpoint cell growth assays were performed by plating a total of 1 x 10^3^ cells per well in a 96-well plate containing RPMI/MOPS media buffered at pH 7 <1% DMSO with or without antifungal drug and incubated at 37°C for 72 hours.

### 5.4 Melanin-Induction Assay

Cryptococcal strains were cultured overnight in YPD liquid media at 30°C shaking for ∼16 hours. Overnight cultures of *Cryptococcus* were washed in Phosphate Buffered Saline (PBS) three times and resuspended in Ultrapure DNase/RNase-Free Distilled Water. Yeast cells were counted and diluted to a concentration of 2 x 10^7^. Of those dilutions, 5 µl (1 x 10^5^ cells per spot) was spotted onto L-DOPA plates in duplicate with at least two biological replicates. L-DOPA plates were incubated at 37°C for 72 hours in ambient air or 5% CO_2_ and imaged by scanning plates on an Epson Expression 10000 XL scanner. After the 72-hour time point, L-DOPA plates were further incubated at 37°C for a total of 10 days and reimaged. In vitro assessment of cryptococcal virulence related to melanin production is commonly performed by growing yeast cells on melanin-inducing L-DOPA media (nutrient limiting media) at the host-like temperature of 37°C in ambient air (50). To assess the effect that exposure to multiple host-like stress conditions have on colony phenotypes and to compare relative virulence by melanin production, we grew strains on melanin-inducing L-DOPA agar and incubated them at 37°C in either ambient air or 5% CO_2_. After 72 hours and 120 hours we scored colony pigment under both air conditions. Because melanin production can also be affected by yeast cell abundance, we modified our assay in an attempt to reduce this affect between strains with varying fitness in CO_2_ by spotting 100,000 yeast cells of each strain on L-DOPA media and incubating the plates at 37°C in ambient air or in 5% CO_2_ followed by visualization at 72 hours and 120 hours. In conjunction with scoring colony pigment on a 5-point scale (0=White, 1=Ivory/Off-White, 2=Light Brown, 3=Brown, and 4=Dark Brown), we also scored colony consistency using the same 4-point scale used in our initial observations of condition-based phenotypic switching. Statistical analyses were performed using GraphPad Prism 9.

### 5.5 Capsule Formation Assay

Cryptococcal strains were cultured overnight in YPD liquid media at 30°C shaking for ∼16 hours. Overnight cultures of *Cryptococcus* were washed and resuspended in PBS. Cells were counted and 4 x 10^5^ cells were added to each well of a 24-well plate that contained DMEM (Gibco) and incubated at 37°C in 5% CO_2_for 72 hours. Cryptococcal yeast cells were harvested, mixed with India Ink, and visualized by microscopy. Images were captured using a ZEISS Axio Imager M2 microscope, camera, software. Quantitative analysis was performed by visually inspecting images in the bright field and yeast cell body and capsule were measured. The capsule phenotypes of viable cells were imaged, characterized, and counted. The data represent the average of at least two biological replicates for which at least 100 yeast cells were counted. Error bars indicate standard deviation. Statistical analyses were performed using GraphPad Prism 9.

### 5.6 Genomic Analysis

To determine if phenotypic heterogeneity represented genotypic variation, multiple phenotypically unique single colony isolates from individual serial patient specimens were collected for subsequent sequencing. For patient specimens that were phenotypically homogenous, one single colony was collected. Colonies were sequenced on the Illumina NextSeq 6000 platform and reads were mapped to a concatenated mega-genome composed of representative reference genomes from each cryptococcal molecular type. Genomes included *C. neoformans* (VNI, serotype A), *C. deneoformans* (VNIV, serotype D), and *C. gattii* (VGI, serotypes B, C). Patient strains that had a majority of reads mapping to more than one reference genome were classified as hybrids. Sites with single nucleotide polymorphisms (SNPs) and insertion/deletions (indels) relative to the reference strain were called using SAMtools. The difference in the number of shared SNP sites, relative to the reference strain, was used as a measure of genetic distance between phenotypically distinct isolates from single patient specimens. Dissemination and persistence of specific cryptococcal strain lineages were identified in clinical samples collected at different body sites and/or different collection times using these criteria.

## Supporting information

Supplemental Table 1 - Patient and Strain Information

Supplemental Table 2 - Antifungal Susceptibility Area Under the Curve

## Data Availability

Genomic Data are available at https://github.com/magwenelab/chronic_crypto_evolution

All other data produced in the present study are available upon reasonable request to the authors.

https://github.com/magwenelab/chronic_crypto_evolution

## Acknowledgements

The research reported in this publication was supported by the National Institute of Allergy and Infectious Diseases of the National Institutes of Health under award numbers: R21AI178058 (JRP, PMM), R01AI 73896 (JRP), R01AI93257 (JRP), R01AI133654 (PMM), T32AI052080 (MCM)

