## Supplemental Table 1 - Patient and Strain Information for "Genome restructuring and lineage diversification of *Cryptococcus neoformans* during chronic infection of human hosts"

| Patient Characteristics | | | | Isolate Information | | | | | | | | Days of Antifungal Exposure† | | | Antifungal Susceptibility | | Cell Measurements | | | | | | | | Colony Viscosity | | | | | Colony Pigment | | Growth | |
|---|---|---|---|---|---|---|---|---|---|---|---|---|---|---|---|---|---|---|---|---|---|---|---|---|---|---|---|---|---|---|---|---|---|
| | | | | | | | | | | | | | | | | | YPD, Air, 30ºC, 72h | | DMEM, 5%CO2, 37ºC, 72h | | | | | | YPD Agar | | | L-DOPA Agar | | L-DOPA Agar | | RPMI/MOPS | |
| | | | | | | | | | | | | | | | 72h | 72h | Whole Cell Diameter | | | | Capsule Thickness | | Cell Body | | 30ºC, 72h | 37ºC, 72h | 37ºC, 72h | 37ºC, 120h | | 37ºC, 120h | | 37ºC, 72h | |
| Pt ID | Sex | Comorbidity | Age Range | DOI | Strain ID | Source | Species | Serotype | Molecular Type | Partial Chr. Deletion | Chr. Dup. | AmB | 5FC | FLC | AmB MIC (µg/ml) | FLC MIC (µg/ml) | Avg. (µm) | SD (µm) | Avg. (µm) | SD (µm) | Avg. (µm) | SD (µm) | Avg. (µm) | SD (µm) | Ambient | | 5%CO2 | Ambient | 5%CO2 | Ambient | 5% CO2 | Avg. OD600nm | SD OD600nm |
| Reference | Male | Hodgkin's Disease | 20-34 | 0 | H99 | CSF | *C. neoformans* | A | VNIb | | | 0 | 0 | 0 | 0.5 | 4 | 7.9 | 1.6 | 11.1 | 1.9 | 2.09 | 0.73 | 6.9 | 0.8 | 1 | 1 | 1 | 1 | 3 | 3 | 0 | 0.543 | 0.035 |
| NC1 | Male | HIV | 20-34 | 20 | MCM 271 - A | Sputum | *C. neoformans* | A | VNIb | | | 5 | 5 | 6 | 0.5 | 2 | 8.2 | 1.7 | 7.2 | 1.7 | 0.69 | 0.60 | 5.8 | 1.0 | 1 | 1 | 2 | 2 | 3 | 1 | 0 | 0.553 | 0.015 |
| | | | | 20 | MCM 271 - E | Sputum | *C. neoformans* | A | VNIb | | | 5 | 5 | 6 | 0.25 | 1 | 9.0 | 2.2 | 12.4 | 3.0 | 2.90 | 0.77 | 6.6 | 1.7 | 2 | 3 | 3 | 3 | 3 | 1 | 0 | 0.523 | 0.016 |
| | | | | 201 | MCM 284 - F | CSF | *C. neoformans* | A | VNIb | 10 | | 5 | 5 | 6 | 0.25 | 8 | 8.5 | 1.7 | 12.2 | 1.2 | 2.79 | 0.49 | 6.6 | 0.5 | 1 | 1 | 1 | 1 | 1 | 3 | 1 | 0.521 | 0.022 |
| | | | 35-44 | 290 | MCM 528 - E | CSF | *C. neoformans* | A | VNIb | | | 12 | 12 | 6 | 0.25 | 4 | 9.8 | 2.4 | 11.6 | 2.3 | 2.37 | 0.68 | 6.8 | 1.2 | 1 | 1 | 1 | 1 | 1 | 3 | 0 | 0.548 | 0.010 |
| | | | | 290 | MCM 528 - F | CSF | *C. neoformans* | A | VNIb | 10 | | 12 | 12 | 6 | 0.25 | 8 | 9.4 | 2.4 | 12.7 | 1.4 | 3.12 | 0.55 | 6.5 | 0.6 | 1 | 1 | 1 | 1 | 1 | 3 | 1 | 0.537 | 0.004 |
| | | | | 379 | MCM 094 - A | CSF | *C. neoformans* | A | VNIb | 10 | | 20 | 20 | 6 | 0.5 | 8 | 9.8 | 3.4 | 12.4 | 1.5 | 3.11 | 0.56 | 6.1 | 0.6 | 1 | 1 | 1 | 1 | 1 | 3 | 1 | 0.476 | 0.030 |
| | | | | 379 | MCM 094 - B | CSF | *C. neoformans* | A | VNIb | 10 | 12 | 20 | 20 | 6 | 0.125 | 4 | 10.5 | 3.1 | 15.2 | 1.6 | 4.16 | 0.73 | 6.9 | 1.2 | 2 | 1 | 1 | 0 | 1 | 3 | 1 | 0.520 | 0.045 |
| | | | | 418 | MCM 095 - A | CSF | *C. neoformans* | A | VNIb | 10 | | 26 | 26 | 7 | 0.5 | 8 | 8.2 | 2.1 | 12.4 | 1.2 | 3.04 | 0.50 | 6.3 | 0.6 | 1 | 1 | 1 | 2 | 1 | 3 | 1 | 0.539 | 0.020 |
| | | | | 418 | MCM 095 - B | CSF | *C. neoformans* | A | VNIb | 10 | | 26 | 26 | 7 | 0.25 | 8 | 8.7 | 2.4 | 12.3 | 2.5 | 3.11 | 1.10 | 6.1 | 0.8 | 1 | 2 | 3 | 3 | 3 | 4 | 1 | 0.580 | 0.010 |
| | | | | 471 | MCM 096 - A | CSF | *C. neoformans* | A | VNIb | 10 | | 31 | 31 | 18 | 0.125 | 4 | 8.8 | 2.6 | 15.1 | 2.5 | 4.21 | 1.03 | 6.6 | 1.0 | 1 | 1 | 1 | 1 | 1 | 3 | 3 | 0.482 | 0.019 |
| | | | | 543 | MCM 101 - B | CSF | *C. neoformans* | A | VNIb | 10 | | 38 | 31 | 29 | 0.25 | 8 | 9.4 | 2.1 | 12.1 | 1.2 | 2.73 | 0.55 | 6.6 | 0.5 | 1 | 1 | 1 | 1 | 1 | 3 | 2 | 0.508 | 0.014 |
| | | | | 543 | MCM 101 - C | CSF | *C. neoformans* | A | VNIb | 10 | 12 | 38 | 31 | 29 | 0.5 | 8 | 9.8 | 2.7 | 10.8 | 1.1 | 2.25 | 0.55 | 6.3 | 0.5 | 1 | 0 | 0 | 1 | 1 | 3 | 2 | 0.580 | 0.011 |
| | | | | 583 | MCM 103 - A | CSF | *C. neoformans* | A | VNIb | 10 | 12* | 43 | 31 | 29 | 0.5 | 8 | 10.2 | 3.1 | 8.7 | 1.6 | 1.23 | 0.81 | 6.3 | 0.7 | 1 | 1 | 1 | 1 | 1 | 3 | 2 | 0.457 | 0.051 |
| | | | | 583 | MCM 103 - B | CSF | *C. neoformans* | A | VNIb | 10 | 1* | 43 | 31 | 29 | 0.5 | 16 | 11.1 | 3.1 | 10.2 | 1.3 | 1.84 | 0.58 | 6.5 | 0.6 | 1 | 1 | 1 | 1 | 1 | 3 | 2 | 0.428 | 0.024 |
| | | | | 605 | MCM 104 - A | CSF | *C. neoformans* | A | VNIb | 10 | | 43 | 31 | 29 | 0.5 | 16 | 9.3 | 2.3 | 8.2 | 2.2 | 1.05 | 0.94 | 6.1 | 0.7 | 1 | 1 | 1 | 1 | 1 | 4 | 2 | 0.437 | 0.009 |
| | | | | 605 | MCM 104 - B | CSF | *C. neoformans* | A | VNIb | 10 | | 43 | 31 | 29 | 0.5 | 16 | 10.5 | 2.8 | 10.3 | 1.5 | 2.06 | 0.73 | 6.2 | 0.5 | 2 | 3 | 3 | 3 | 3 | 3 | 2 | 0.478 | 0.042 |
| | | | | 605 | MCM 104 - C | CSF | *C. neoformans* | A | VNIb | 10 | | 43 | 31 | 29 | 0.5 | 16 | 9.9 | 2.9 | X | X | X | X | X | X | 1 | 1 | 3 | 3 | 3 | 3 | 2 | 0.476 | 0.022 |
| | | | | 648 | MCM 107 - A | CSF | *C. neoformans* | A | VNIb | 10 | | 48 | 35 | 29 | 0.5 | 16 | 9.8 | 2.5 | 7.5 | 1.0 | 0.59 | 0.33 | 6.3 | 0.8 | 1 | 2 | 3 | 3 | 3 | 4 | 1 | 0.468 | 0.035 |
| | | | | 648 | MCM 107 - B | CSF | *C. neoformans* | A | VNIb | 10 | | 48 | 35 | 29 | 0.5 | 16 | 10.0 | 2.7 | 7.5 | 1.1 | 0.64 | 0.42 | 6.2 | 0.7 | 1 | 2 | 3 | 3 | 3 | 3 | 3 | 0.448 | 0.007 |
| | | | | 648 | MCM 107 - J | CSF | *C. neoformans* | A | VNIb | 10 | 12 | 48 | 35 | 29 | 0.5 | 8 | 10.3 | 3.3 | 6.8 | 0.9 | 0.28 | 0.26 | 6.3 | 0.7 | 1 | 1 | 1 | 1 | 1 | 4 | 4 | 0.486 | 0.012 |
| | | | | 648 | MCM 107 - K | CSF | *C. neoformans* | A | VNIb | 10 | | 48 | 35 | 29 | 0.5 | 16 | 8.2 | 2.1 | 7.7 | 1.2 | 0.69 | 0.52 | 6.3 | 0.6 | 1 | 1 | 3 | 3 | 3 | 3 | 2 | 0.491 | 0.034 |
| | | | | 724 | MCM 297 - A | CSF | *C. neoformans* | A | VNIb | 10 | 1 | 48 | 35 | 29 | 0.5 | 32 | 9.4 | 3.7 | X | X | X | X | X | X | 1 | 2 | 2 | 3 | 2 | 4 | 4 | 0.531 | 0.011 |
| | | | | 724 | MCM 297 - C | CSF | *C. neoformans* | A | VNIb | 10 | 12 | 48 | 35 | 29 | 0.5 | 8 | 8.0 | 2.4 | 8.5 | 1.1 | 1.02 | 0.50 | 6.5 | 0.5 | 1 | 2 | 2 | 3 | 3 | 4 | 3 | 0.605 | 0.017 |
| | | | | 724 | MCM 297 - D | CSF | *C. neoformans* | A | VNIb | 10 | 6 | 48 | 35 | 29 | 0.5 | 8 | 7.8 | 2.2 | 8.6 | 1.2 | 1.06 | 0.53 | 6.5 | 0.6 | 1 | 2 | 3 | 3 | 3 | 3 | 1 | 0.546 | 0.015 |
| | | | | 774 | MCM 298 - A | CSF | *C. neoformans* | A | VNIb | 10 | 12 | 55 | 42 | 29 | 0.5 | 8 | 7.8 | 2.2 | 7.5 | 0.8 | 0.62 | 0.26 | 6.3 | 0.6 | 1 | 1 | 2 | 2 | 3 | 4 | 3 | 0.577 | 0.025 |
| | | | | 774 | MCM 298 - J | CSF | *C. neoformans* | A | VNIb | 10 | 1 | 55 | 42 | 29 | 0.5 | 16 | 10.1 | 2.9 | 8.7 | 1.4 | 0.63 | 0.41 | 7.4 | 1.1 | 1 | 1 | 2 | 3 | 2 | 4 | 3 | 0.551 | 0.006 |
| | | | | 855 | MCM 300 - A | CSF | *C. neoformans* | A | VNIb | 10 | 12 | 68 | 53 | 29 | 0.5 | 8 | 10.3 | 2.5 | 7.2 | 1.1 | 0.62 | 0.32 | 6.0 | 0.6 | 1 | 2 | 2 | 3 | 3 | 4 | 3 | 0.543 | 0.014 |
| | | | | 855 | MCM 300 - D | CSF | *C. neoformans* | A | VNIb | 10 | 1 | 68 | 53 | 29 | 0.5 | 16 | 9.7 | 3.7 | X | X | X | X | X | X | 1 | 1 | 2 | 2 | 1 | 3 | 4 | 0.536 | 0.010 |
| | | | | 855 | MCM 300 - K | CSF | *C. neoformans* | A | VNIb | 10 | 12 | 68 | 53 | 29 | 0.5 | 8 | 10.2 | 2.6 | 10.2 | 1.6 | 1.65 | 0.77 | 6.9 | 0.6 | 1 | 2 | 2 | 2 | 2 | 4 | 3 | 0.556 | 0.030 |
| | | | | 855 | MCM 300 - L | CSF | *C. neoformans* | A | VNIb | 10 | 1 | 68 | 53 | 29 | 0.5 | 16 | 9.4 | 3.4 | X | X | X | X | X | X | 1 | 2 | 2 | 2 | 1 | 4 | 4 | 0.528 | 0.011 |
| NC6 | Male | HIV | 35-44 | 3 | MCM 529 - B | CSF | *C. neoformans* | A | VNIb | | | 0 | 0 | 2 | 0.25 | 4 | 10.2 | 2.8 | X | X | X | X | X | X | 2 | 2 | 2 | 2 | 3 | 4 | 0 | 0.416 | 0.017 |
| | | | | 3 | MCM 529 - A | CSF | *C. neoformans* | A | VNIb | | | 0 | 0 | 2 | 0.25 | 4 | 9.7 | 3.2 | X | X | X | X | X | X | 2 | 2 | 2 | 2 | 3 | 4 | 0 | 0.430 | 0.016 |
| | | | 55-64 | 7619 | MCM 012 | CSF | *C. neoformans* | A | VNIa | | 5 | 9 | 9 | 2427 | 0.25 | 4 | 9.4 | 3.1 | X | X | X | X | X | X | 1 | 2 | 2 | 1 | 3 | 1 | 1 | 0.444 | 0.012 |
| NC8 | Male | HIV | 20-34 | 0 | MCM 109 - A | CSF | *C. neoformans x C. deneoformans* | AD Hybrid | VNI/VNIV | | | 0 | 0 | 0 | 0.25 | 16 | 10.6 | 3.1 | 8.5 | 1.2 | 0.2 | 0.2 | 8.0 | 1.0 | 1 | 1 | 1 | 1 | 1 | 2 | 2 | 0.459 | 0.017 |
| | | | | 0 | MCM 109 - B | CSF | *C. neoformans x C. deneoformans* | AD Hybrid | VNI/VNIV | | | 0 | 0 | 0 | 0.25 | 16 | 13.2 | 3.9 | 10.3 | 1.7 | 1.3 | 0.4 | 7.8 | 1.2 | 1 | 1 | 1 | 1 | 1 | 2 | 2 | 0.442 | 0.023 |
| | | | | 0 | MCM 109 - C | CSF | *C. neoformans x C. deneoformans* | AD Hybrid | VNI/VNIV | | | 0 | 0 | 0 | 0.5 | 16 | 12.2 | 3.6 | 11.1 | 1.8 | 1.5 | 0.5 | 8.0 | 1.2 | 1 | 1 | 1 | 1 | 1 | 2 | 2 | 0.457 | 0.026 |
| | | | 35-44 | 210 | MCM 112 | CSF | *C. neoformans x C. deneoformans* | AD Hybrid | VNI/VNIV | | | 34 | 33 | 115 | 0.25 | 16 | 10.4 | 3.0 | 8.8 | 1.1 | 0.5 | 0.2 | 7.9 | 1.1 | 1 | 1 | 1 | 1 | 1 | 2 | 2 | 0.444 | 0.014 |
| | | | | 256 | MCM 113 - A | CSF | *C. neoformans x C. deneoformans* | AD Hybrid | VNI/VNIV | | | 48 | 33 | 115 | 0.25 | 16 | 10.4 | 3.0 | 9.1 | 1.7 | 0.9 | 0.5 | 7.4 | 1.2 | 1 | 1 | 1 | 1 | 1 | 2 | 2 | 0.441 | 0.032 |
| | | | | 256 | MCM 113 - B | CSF | *C. neoformans x C. deneoformans* | AD Hybrid | VNI/VNIV | | | 48 | 33 | 115 | 0.25 | 16 | 9.8 | 2.7 | 9.8 | 1.6 | 0.9 | 0.4 | 7.9 | 1.2 | 1 | 1 | 1 | 1 | 1 | 2 | 2 | 0.397 | 0.027 |
| | | | | 256 | MCM 113 - C | CSF | *C. neoformans x C. deneoformans* | AD Hybrid | VNI/VNIV | | | 48 | 33 | 115 | 0.25 | 32 | 8.5 | 2.3 | 8.7 | 1.2 | 0.4 | 0.2 | 7.9 | 1.1 | 1 | 1 | 1 | 1 | 1 | 2 | 2 | 0.449 | 0.022 |
| | | | | 319 | MCM 119 | CSF | *C. neoformans x C. deneoformans* | AD Hybrid | VNI/VNIV | | | 54 | 36 | 166 | 0.25 | 32 | 9.4 | 2.9 | 10.8 | 2.0 | 1.5 | 0.6 | 7.9 | 1.2 | 1 | 0 | 0 | 0 | 1 | 2 | 2 | 0.450 | 0.018 |
| | | | | 392 | MCM 125 - A | CSF | *C. albidus* | | | | | 71 | 52 | 166 | 0.5 | > 128 | 11.0 | 3.9 | 5.9 | 1.0 | 0.3 | 0.2 | 5.3 | 0.9 | 2 | 2 | 2 | 1 | 1 | 1 | 0 | 0.438 | 0.028 |
| | | | | 392 | MCM 125 - B | CSF | *C. neoformans x C. deneoformans* | AD Hybrid | VNI/VNIV | | | 71 | 52 | 166 | 0.25 | 32 | 12.4 | 3.9 | 11.2 | 2.0 | 1.5 | 0.5 | 8.3 | 1.4 | 1 | 1 | 1 | 1 | 1 | 2 | 1 | 0.432 | 0.016 |
| | | | | 392 | MCM 125 - C | CSF | *C. neoformans x C. deneoformans* | AD Hybrid | VNI/VNIV | | | 71 | 52 | 166 | 0.25 | 64 | 9.9 | 2.5 | 10.9 | 1.8 | 1.5 | 0.5 | 8.0 | 1.3 | 1 | 1 | 1 | 1 | 1 | 2 | 1 | 0.381 | 0.015 |
| NC83 | Male | HIV | 35-44 | 0 | MCM 299 - B | CSF | *C. neoformans* | A | VNII | | | 0 | 0 | 0 | 0.125 | 1 | 8.1 | 1.8 | X | X | X | X | X | X | 1 | 3 | 3 | 1 | 2 | 2 | 1 | 0.456 | 0.017 |
| | | | | 0 | MCM 299 - J | CSF | *C. neoformans* | A | VNII | | | 0 | 0 | 0 | 0.125 | 1 | 11.4 | 3.7 | X | X | X | X | X | X | 0 | 2 | 2 | 1 | 2 | 1 | 0 | 0.502 | 0.011 |
| | | | 45-64 | 3631 | MCM 530 - A | CSF | *C. neoformans* | A | VNIb | | | 23 | 15 | 35 | 0.5 | 1 | 9.2 | 2.5 | X | X | X | X | X | X | 1 | 2 | 1 | 1 | 3 | 2 | 0 | 0.440 | 0.014 |
| | | | | 3631 | MCM 530 - F | CSF | *C. neoformans* | A | VNIb | | | 23 | 15 | 35 | 0.25 | 0.25 | 10.1 | 2.9 | X | X | X | X | X | X | 1 | 2 | 1 | 1 | 2 | 2 | 0 | 0.443 | 0.017 |
| NC94 | Male | HIV | 35-44 | 0 | MCM 136 - A | Blood | *C. neoformans* | A | VNII/VNBII | | | 0 | 0 | 139 | 0.25 | 2 | 10.3 | 2.7 | 13.0 | 1.9 | 2.8 | 0.6 | 7.4 | 0.9 | 1 | 1 | 1 | 1 | 1 | 1 | 0 | 0.406 | 0.016 |
| | | | | 735 | MCM 450 - A | Blood | *C. neoformans* | A | VNII/VNBII | | | 2 | 0 | 542 | 0.125 | 2 | 10.8 | 3.9 | 9.4 | 1.7 | 1.1 | 0.5 | 7.3 | 1.2 | 1 | 0 | 0 | 1 | 0 | 1 | 1 | 0.362 | 0.016 |
| | | | | 735 | MCM 450 - N | Blood | *C. neoformans* | A | VNII/VNBII | | | 2 | 0 | 542 | 0.125 | 2 | 10.7 | 2.8 | 11.3 | 1.8 | 1.5 | 0.4 | 8.2 | 1.3 | 1 | 1 | 1 | 1 | 1 | 2 | 1 | 0.372 | 0.009 |
| | | | | 735 | MCM 531 - A | CSF | *C. neoformans* | A | VNII/VNBII | | | 2 | 0 | 542 | 0.25 | 2 | 10.3 | 2.8 | 10.5 | 1.5 | 1.7 | 0.4 | 7.1 | 1.0 | 2 | 1 | 1 | 1 | 2 | 1 | 0 | 0.434 | 0.022 |
| | | | | 735 | MCM 531 - C | CSF | *C. neoformans* | A | VNII/VNBII | | | 2 | 0 | 542 | 0.125 | 4 | 9.8 | 2.6 | 11.4 | 1.7 | 2.1 | 0.5 | 7.2 | 0.9 | 2 | 1 | 1 | 1 | 1 | 1 | 0 | 0.463 | 0.030 |
| NC183 | Female | SOT | 20-34 | 0 | MCM 197 - G | Sputum | *C. neoformans* | A | VNIa | | | 0 | 0 | 0 | 0.25 | 8 | 8.3 | 2.1 | X | X | X | X | X | X | 1 | 1 | 1 | 1 | 3 | 2 | 1 | 0.615 | 0.011 |
| | | | | 0 | MCM 197 - H | Sputum | *C. neoformans* | A | VNIa | | | 0 | 0 | 0 | 0.25 | 4 | 9.4 | 2.3 | X | X | X | X | X | X | 0 | 1 | 1 | 1 | 3 | 1 | 1 | 0.608 | 0.008 |
| | | | | 445 | MCM 100 - B | Lung | *C. neoformans* | A | VNIa | | | 0 | 0 | 128 | 0.5 | 8 | 10.1 | 2.8 | X | X | X | X | X | X | 0 | 1 | 1 | 1 | 2 | 3 | 2 | 0.473 | 0.018 |
| | | | | 445 | MCM 100 - C | Lung | *C. neoformans* | A | VNIa | | | 0 | 0 | 128 | 0.5 | 8 | 10.1 | 3.4 | X | X | X | X | X | X | 0 | 0 | 0 | 1 | 2 | 3 | 2 | 0.492 | 0.058 |

†Cumulative days of antifungal exposure prior to day of a positive *Cryptococcus* culture.

*Partial chromosomal duplication

Colony Viscosity: 0 - Dry, 1 - Creamy, 2 - Shiny, 3 - Mucoid

Colony Melanin Pigment: 0 - White, 1 - Ivory, 2 - Light Brown, 3 - Brown, 4 - Dark Brown

X Indicates measurement was not completed
