## Supplemental Table 2 - Antifungal Susceptibility Area Under the Curve for "Genome restructuring and lineage diversification of *Cryptococcus neoformans* during chronic infection of human hosts"

| Supplementary Table 2 | | | | | Fluconazole (0 - 16 ug/ml) | | | Amphotericin B (0 - 0.5 ug/ml) | | |
|---|---|---|---|---|---|---|---|---|---|---|
| Patient | DOI | Partial Chr. Deletion | Chr. Dup. | Strain ID | Total Area | Std. Error | 95% Confidence Interval | Total Area | Std. Error | 95% Confidence Interval |
| NC1 | 20 | | | MCM 271A | 3.104 | 0.0968 | 2.914 to 3.293 | 0.1433 | 0.01629 | 0.1113 to 0.1752 |
| NC1 | 20 | | | MCM 271E | 3.011 | 0.05201 | 2.909 to 3.113 | 0.1172 | 0.00158 | 0.1141 to 0.1203 |
| NC1 | 201 | 10 | | MCM 284F | 3.689 | 0.07094 | 3.550 to 3.828 | 0.1363 | 0.00161 | 0.1331 to 0.1394 |
| NC1 | 290 | | | MCM 528E | 3.059 | 0.04745 | 2.966 to 3.152 | 0.1157 | 0.001623 | 0.1125 to 0.1189 |
| NC1 | 290 | 10 | | MCM 528F | 3.836 | 0.09687 | 3.646 to 4.025 | 0.1282 | 0.001406 | 0.1254 to 0.1309 |
| NC1 | 379 | 10 | | MCM 94A | 3.832 | 0.05936 | 3.715 to 3.948 | 0.1706 | 0.0147 | 0.1418 to 0.1994 |
| NC1 | 379 | 10 | 12 | MCM 94B | 3.69 | 0.06478 | 3.563 to 3.817 | 0.1333 | 0.001613 | 0.1301 to 0.1364 |
| NC1 | 418 | 10 | | MCM 95A | 3.872 | 0.09895 | 3.678 to 4.066 | 0.1654 | 0.01769 | 0.1307 to 0.2000 |
| NC1 | 418 | 10 | | MCM 95B | 4.031 | 0.1691 | 3.699 to 4.362 | 0.1323 | 0.003766 | 0.1249 to 0.1397 |
| NC1 | 471 | 10 | | MCM 96A | 3.955 | 0.04279 | 3.871 to 4.039 | 0.1436 | 0.005302 | 0.1332 to 0.1540 |
| NC1 | 543 | 10 | | MCM 101B | 3.971 | 0.04831 | 3.876 to 4.066 | 0.1365 | 0.001909 | 0.1327 to 0.1402 |
| NC1 | 543 | 10 | 12 | MCM 101C | 3.774 | 0.09505 | 3.587 to 3.960 | 0.1673 | 0.0193 | 0.1295 to 0.2051 |
| NC1 | 583 | 10 | 12* | MCM 103A | 4.215 | 0.07421 | 4.069 to 4.360 | 0.1673 | 0.01118 | 0.1454 to 0.1892 |
| NC1 | 583 | 10 | 1* | MCM 103B | 3.941 | 0.03702 | 3.869 to 4.014 | 0.1287 | 0.002451 | 0.1239 to 0.1335 |
| NC1 | 605 | 10 | | MCM 104A | 4.081 | 0.08253 | 3.919 to 4.243 | 0.1378 | 0.00347 | 0.1310 to 0.1446 |
| NC1 | 605 | 10 | | MCM 104B | 3.94 | 0.09527 | 3.753 to 4.127 | 0.1623 | 0.01478 | 0.1334 to 0.1913 |
| NC1 | 605 | 10 | | MCM 104C | 3.917 | 0.06563 | 3.789 to 4.046 | 0.1278 | 0.002162 | 0.1236 to 0.1321 |
| NC1 | 648 | 10 | | MCM 107A | 3.918 | 0.1305 | 3.662 to 4.173 | 0.1627 | 0.0106 | 0.1419 to 0.1835 |
| NC1 | 648 | 10 | | MCM 107B | 3.76 | 0.03711 | 3.687 to 3.833 | 0.1422 | 0.001347 | 0.1396 to 0.1449 |
| NC1 | 648 | 10 | 12 | MCM 107J | 3.432 | 0.08247 | 3.270 to 3.593 | 0.1402 | 0.001937 | 0.1364 to 0.1440 |
| NC1 | 648 | 10 | | MCM 107K | 3.795 | 0.1184 | 3.563 to 4.027 | 0.1623 | 0.01529 | 0.1323 to 0.1923 |
| NC1 | 724 | 10 | 1 | MCM 297A | 4.967 | 0.04428 | 4.880 to 5.054 | 0.1533 | 0.002447 | 0.1485 to 0.1581 |
| NC1 | 724 | 10 | 12 | MCM 297C | 4.163 | 0.1067 | 3.954 to 4.372 | 0.1997 | 0.01357 | 0.1731 to 0.2263 |
| NC1 | 724 | 10 | 6 | MCM 297D | 4.003 | 0.03821 | 3.928 to 4.077 | 0.1528 | 0.00521 | 0.1426 to 0.1630 |
| NC1 | 774 | 10 | 12 | MCM 298A | 4.108 | 0.06299 | 3.985 to 4.231 | 0.1715 | 0.002571 | 0.1665 to 0.1765 |
| NC1 | 774 | 10 | 1 | MCM 298J | 5.438 | 0.2034 | 5.039 to 5.836 | 0.1835 | 0.01186 | 0.1603 to 0.2068 |
| NC1 | 855 | 10 | 12 | MCM 300A | 3.853 | 0.09331 | 3.670 to 4.036 | 0.1633 | 0.004572 | 0.1543 to 0.1723 |
| NC1 | 855 | 10 | 1 | MCM 300D | 4.87 | 0.0996 | 4.675 to 5.065 | 0.1627 | 0.001276 | 0.1602 to 0.1652 |
| NC1 | 855 | 10 | 12 | MCM 300K | 3.983 | 0.106 | 3.775 to 4.190 | 0.1818 | 0.01505 | 0.1523 to 0.2113 |
| NC1 | 855 | 10 | 1 | MCM 300L | 4.844 | 0.07921 | 4.689 to 4.999 | 0.1422 | 0.003284 | 0.1358 to 0.1486 |
| NC6 | 3 | | | MCM 529B | 2.978 | 0.07565 | 2.830 to 3.126 | 0.114 | 0.001741 | 0.1106 to 0.1174 |
| NC6 | 3 | | | MCM 529A | 3.228 | 0.07971 | 3.071 to 3.384 | 0.1163 | 0.002781 | 0.1109 to 0.1218 |
| NC6 | 7619 | | 5 | MCM 12 | 3.365 | 0.07996 | 3.208 to 3.522 | 0.1294 | 0.006682 | 0.1163 to 0.1424 |
| NC8 | 0 | | | MCM 109A | 3.562 | 0.1164 | 3.334 to 3.790 | 0.1143 | 0.002435 | 0.1095 to 0.1190 |
| NC8 | 0 | | | MCM 109B | 3.754 | 0.1224 | 3.514 to 3.994 | 0.1123 | 0.00278 | 0.1069 to 0.1178 |
| NC8 | 0 | | | MCM 109C | 3.609 | 0.05716 | 3.497 to 3.721 | 0.1237 | 0.005555 | 0.1128 to 0.1346 |
| NC8 | 210 | | | MCM 112 | 3.615 | 0.1055 | 3.408 to 3.821 | 0.1037 | 0.001783 | 0.1002 to 0.1072 |
| NC8 | 256 | | | MCM 113A | 3.658 | 0.1192 | 3.424 to 3.891 | 0.1096 | 0.002144 | 0.1054 to 0.1138 |
| NC8 | 256 | | | MCM 113B | 3.401 | 0.07953 | 3.245 to 3.557 | 0.1124 | 0.005684 | 0.1013 to 0.1235 |
| NC8 | 256 | | | MCM 113C | 4.056 | 0.04528 | 3.967 to 4.145 | 0.1076 | 0.001261 | 0.1052 to 0.1101 |
| NC8 | 319 | | | MCM 119 | 4.177 | 0.1416 | 3.900 to 4.455 | 0.1184 | 0.002555 | 0.1134 to 0.1234 |
| NC8 | 392 | | | MCM 125A | 3.45 | 0.07849 | 3.296 to 3.604 | 0.1718 | 0.009144 | 0.1539 to 0.1898 |
| NC8 | 392 | | | MCM 125B | 4.395 | 0.1182 | 4.163 to 4.626 | 0.1214 | 0.004008 | 0.1135 to 0.1293 |
| NC8 | 392 | | | MCM 125C | 4.148 | 0.09561 | 3.961 to 4.335 | 0.1201 | 0.003675 | 0.1129 to 0.1273 |
| NC83 | 0 | | | MCM 299B | 3.369 | 0.03492 | 3.300 to 3.437 | 0.09829 | 0.002055 | 0.09427 to 0.1023 |
| NC83 | 0 | | | MCM 299J | 3.263 | 0.05151 | 3.162 to 3.364 | 0.11 | 0.00142 | 0.1073 to 0.1128 |
| NC83 | 3631 | | | MCM 530A | 3.061 | 0.06587 | 2.932 to 3.190 | 0.1384 | 0.01072 | 0.1174 to 0.1594 |
| NC83 | 3631 | | | MCM 530F | 2.917 | 0.03083 | 2.857 to 2.978 | 0.1201 | 0.00263 | 0.1149 to 0.1252 |
| NC94 | 0 | | | MCM 136A | 3.262 | 0.04461 | 3.175 to 3.350 | 0.1097 | 0.006463 | 0.09701 to 0.1223 |
| NC94 | 735 | | | MCM 450A | 3.146 | 0.03473 | 3.078 to 3.214 | 0.09224 | 0.001422 | 0.08945 to 0.09503 |
| NC94 | 735 | | | MCM 450N | 3.118 | 0.07547 | 2.970 to 3.266 | 0.1023 | 0.002339 | 0.09772 to 0.1069 |
| NC94 | 735 | | | MCM 531A | 3.257 | 0.06218 | 3.135 to 3.378 | 0.1321 | 0.005513 | 0.1213 to 0.1429 |
| NC94 | 735 | | | MCM 531C | 3.233 | 0.03511 | 3.164 to 3.302 | 0.1095 | 0.001611 | 0.1064 to 0.1127 |
| NC183 | 0 | | | MCM 197G | 3.61 | 0.09693 | 3.420 to 3.800 | 0.1585 | 0.01834 | 0.1226 to 0.1944 |
| NC183 | 0 | | | MCM 197H | 3.318 | 0.03076 | 3.258 to 3.378 | 0.132 | 0.00393 | 0.1243 to 0.1397 |
| NC183 | 445 | | | MCM 100B | 3.589 | 0.07358 | 3.444 to 3.733 | 0.1488 | 0.001762 | 0.1454 to 0.1523 |
| NC183 | 445 | | | MCM 100C | 3.791 | 0.1285 | 3.539 to 4.043 | 0.1723 | 0.008893 | 0.1548 to 0.1897 |
