## Supplemental Figures for "Genome restructuring and lineage diversification of *Cryptococcus neoformans* during chronic infection of human hosts"

A.

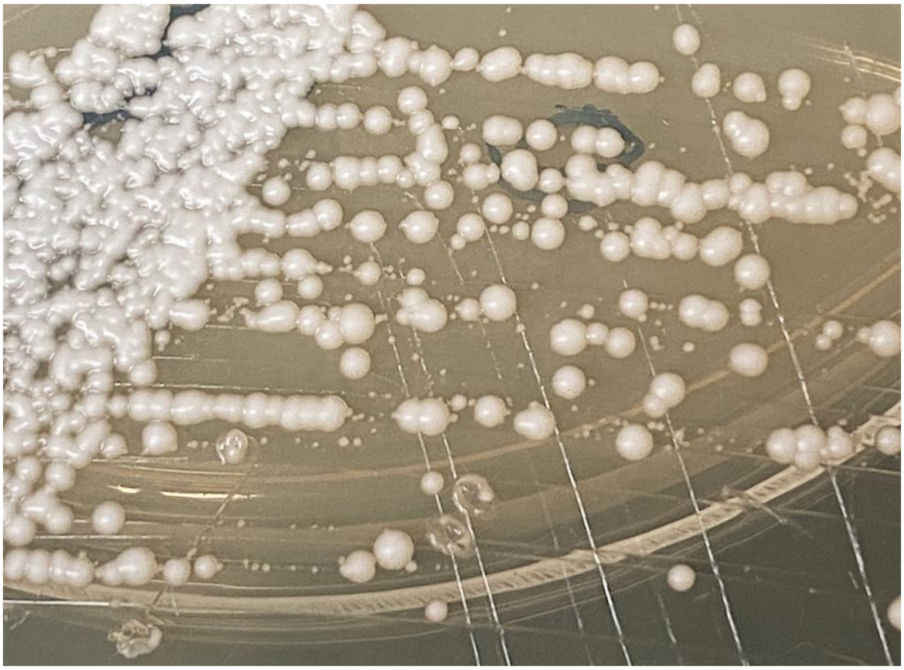

B.

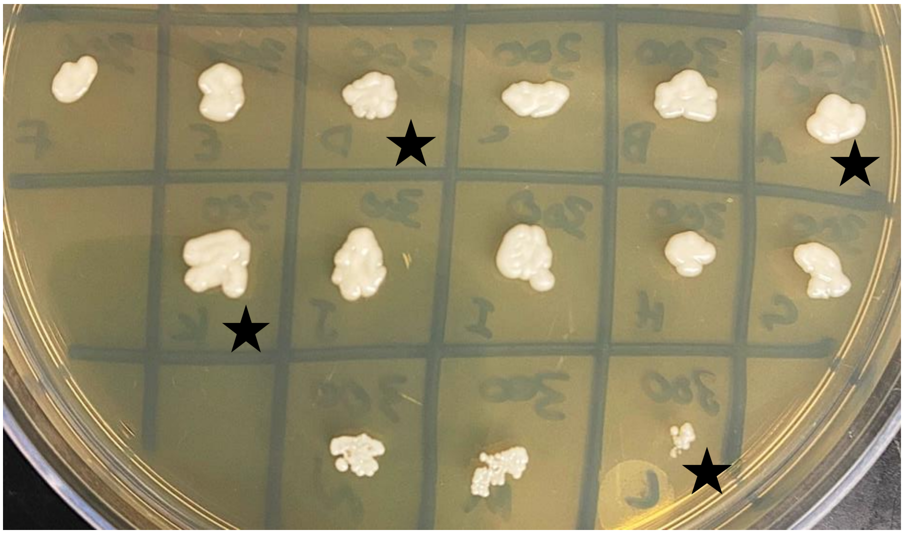

C.

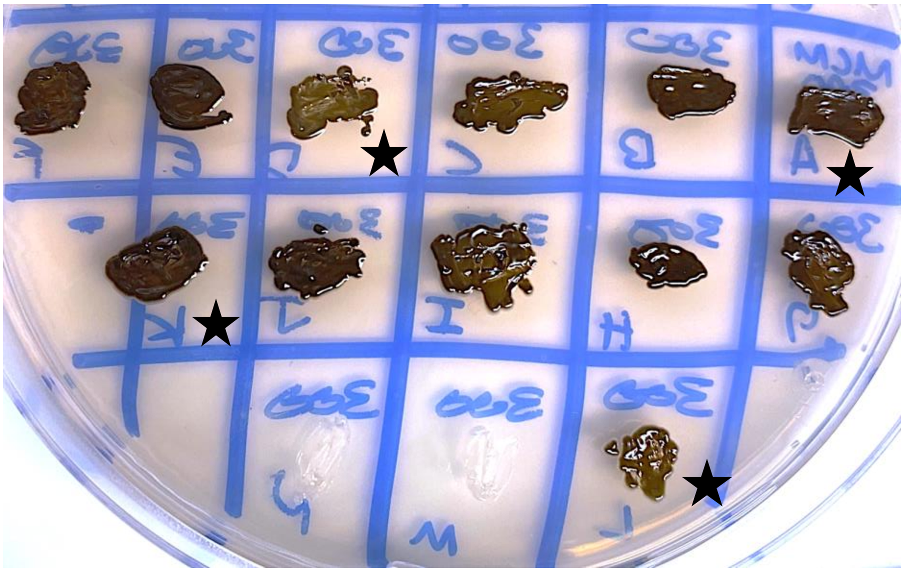

**Figure S1. Example of phenotypic variation within a single patient specimen.** MCM300 is a CSF specimen from patient NC1 collected on day of infection 855. Black stars indicate colonies that were used for further phenotypic analysis and whole genome sequencing.

**A)** MCM300 was streaked from the clinical stock on YPD agar and incubated at 30°C in ambient air for 48-72 hours. Colonies showed phenotypic variation in colony size and colony viscosity. From this plate, 14 phenotypically diverse colonies labeled A through N were picked and patched on YPD agar and L-DOPA agar and incubated at 30°C or 37°C for 72 hours, respectively. **B)** Phenotypic heterogeneity among the patched colonies can be observed in the slight differences of colony viscosity via the presence of both creamy and more mucoid colonies. **C)** Phenotypic variation among MCM300 colonies can be more easily observed when looking at their melanin pigment where MCM300A and and 300K were dark brown and MCM300D and 300L had a light brown pigment.

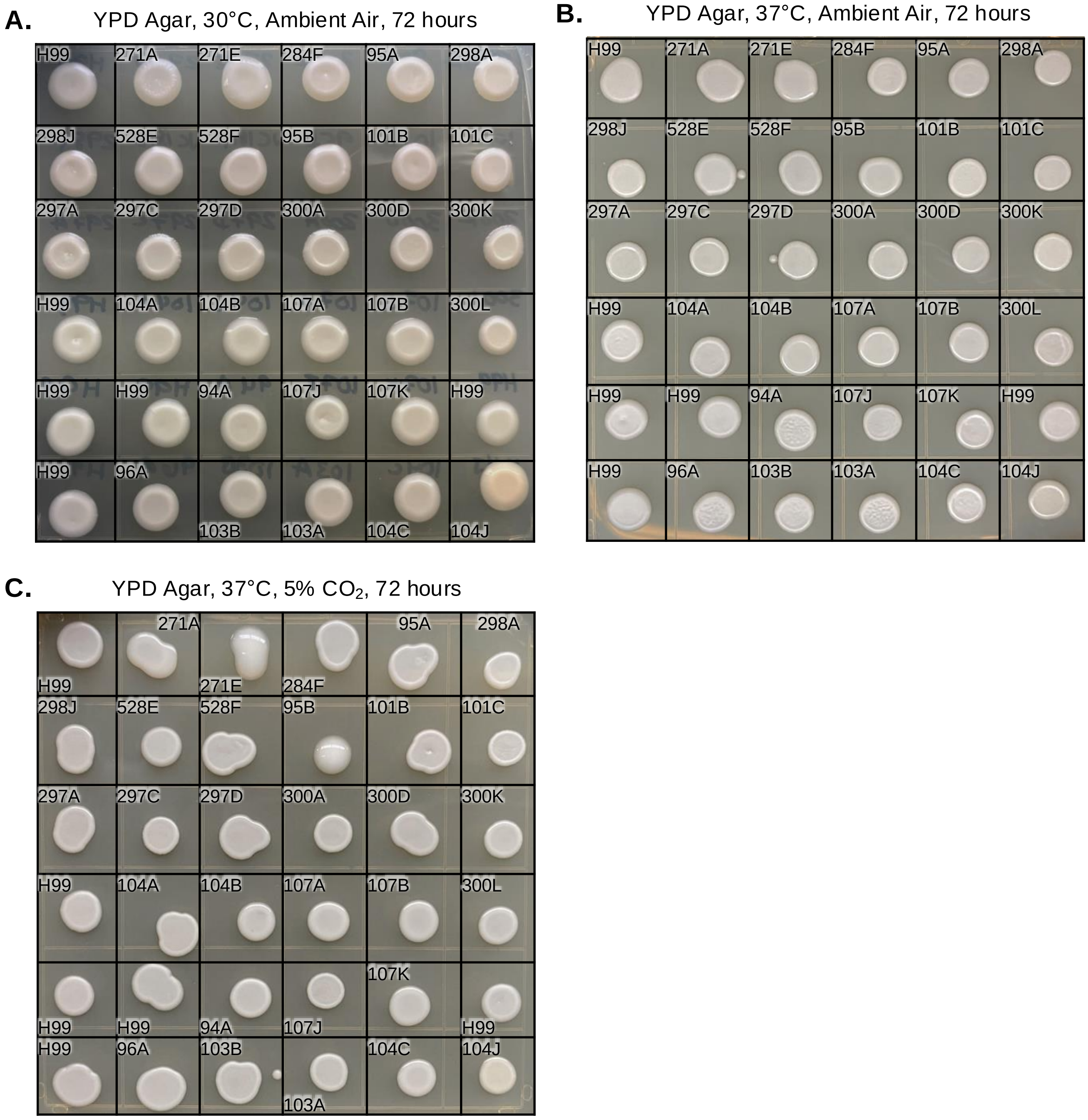


**Figure S2. Phenotypic variation and stress-dependent phenotypic switching on YPD agar is observed among the colonies isolated from serial isolates from patient NC1. A-C)** When grown on YPD agar, differences in colony viscosity can be observed between the cryptococcal strains isolated from serial specimens. Phenotypic switching in colony complexity and colony viscosity can be observed between the different culturing conditions and increasing host-like stressors. Strain MCM104J was identified as *Cryptococcus albidus* indicating that the CSF specimen MCM104 collected on DOI 605 represented a timepoint where patient NC1 experienced a mixed-cryptococcal species infection. H99 was used as a control strain.

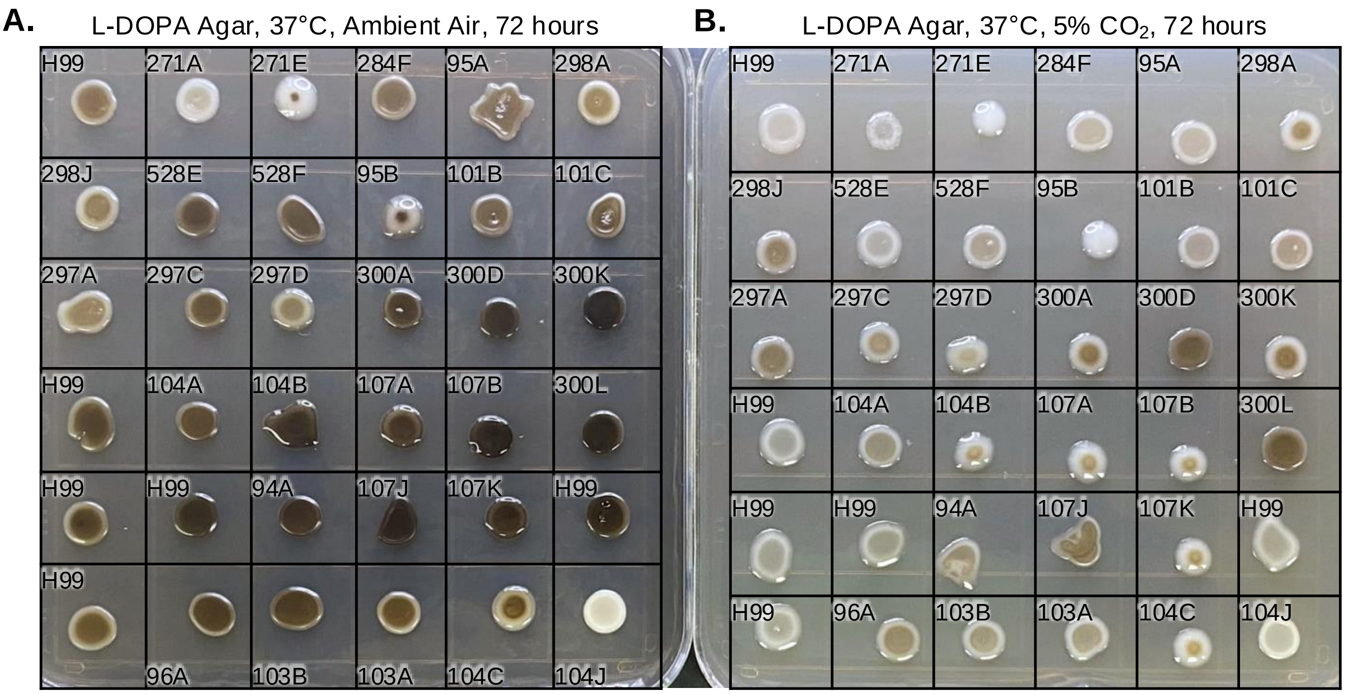


**Figure S3. Phenotypic variation and stress-dependent phenotypic switching on melanin-inducing L-DOPA agar is observed among the colonies isolated from serial isolates from patient NC1.**
**A-B)** When grown on melanin-inducing L-DOPA media, differences in colony pigment and viscosity can be observed between the cryptococcal strains isolated from serial specimens. Phenotypic switching in degree of colony viscosity can be observed between the different culturing conditions with increasing host-like stressors. In addition, a $CO_2$-dependent delay in melanin formation is observed among all strains. When grown Strain MCM104J was identified as *Cryptococcus albidus* indicating that the CSF specimen MCM104 collected on DOI 605 represented a timepoint where patient NC1 experienced a mixed-cryptococcal species infection. H99 was used as a control strain.

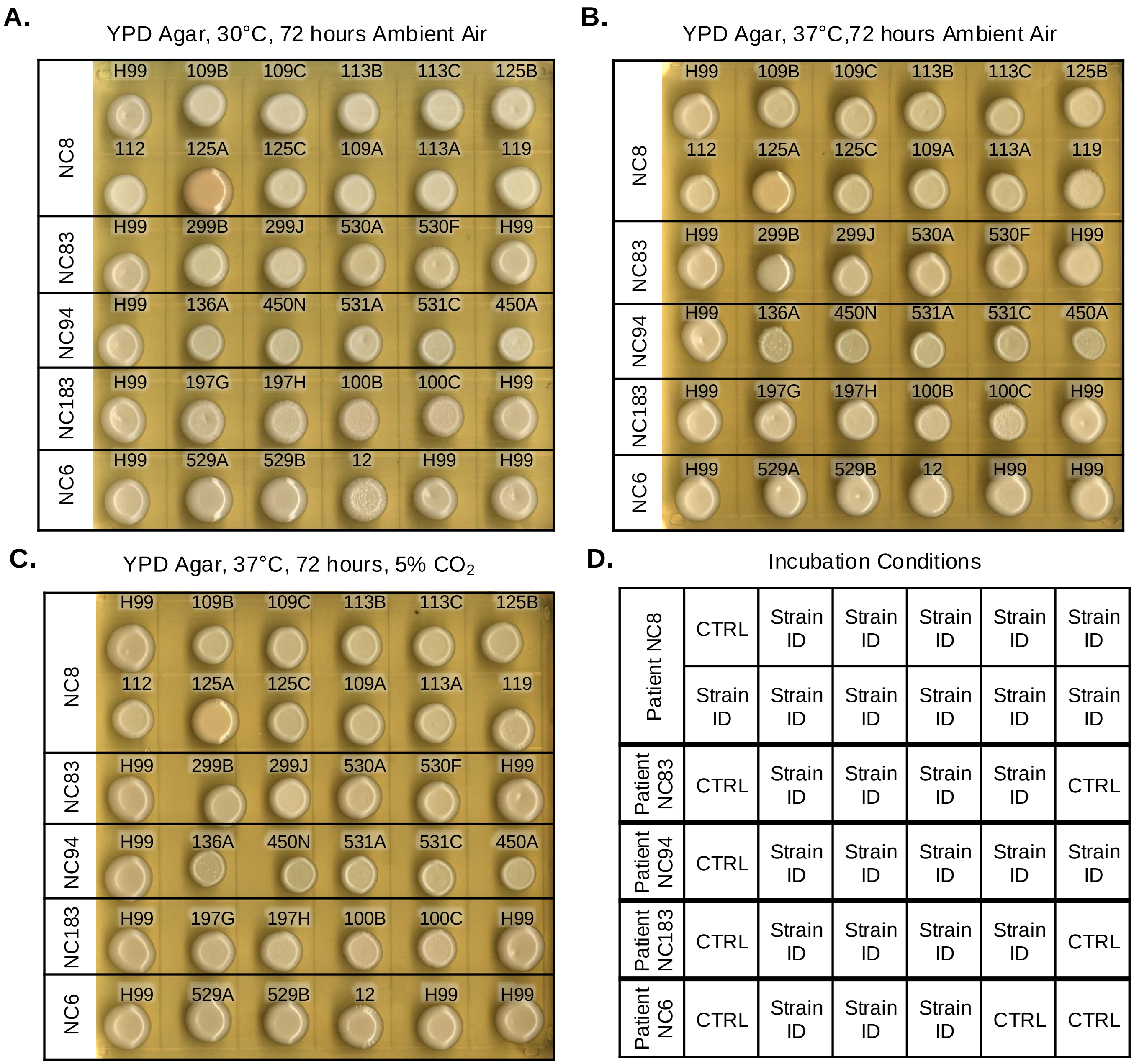


**Figure S4. Phenotypic variation and stress-dependent phenotypic switching on YPD agar is observed among the colonies isolated from serial isolates from patients NC8, NC83, NC94, NC183, and NC6. A-C)** Differences in colony complexity is observed between the colonies isolated from serial specimens of each patient. Phenotypic switching with regard to colony complexity is also observed among isolates from at least one colony from each patient. **D)** Plate map of patient strains where the top two rows are patient NC8 strains, the third row NC83, fourth row NC94, fifth row NC183, and the bottom row is NC6 strains. CTRL – control strain H99. MCM125A was identified to be *Cryptococcus albidus* indicating that the CSF specimen MCM125 represented a timepoint in which patient NC8 had a mixed-cryptococcal species infection.

**E.**

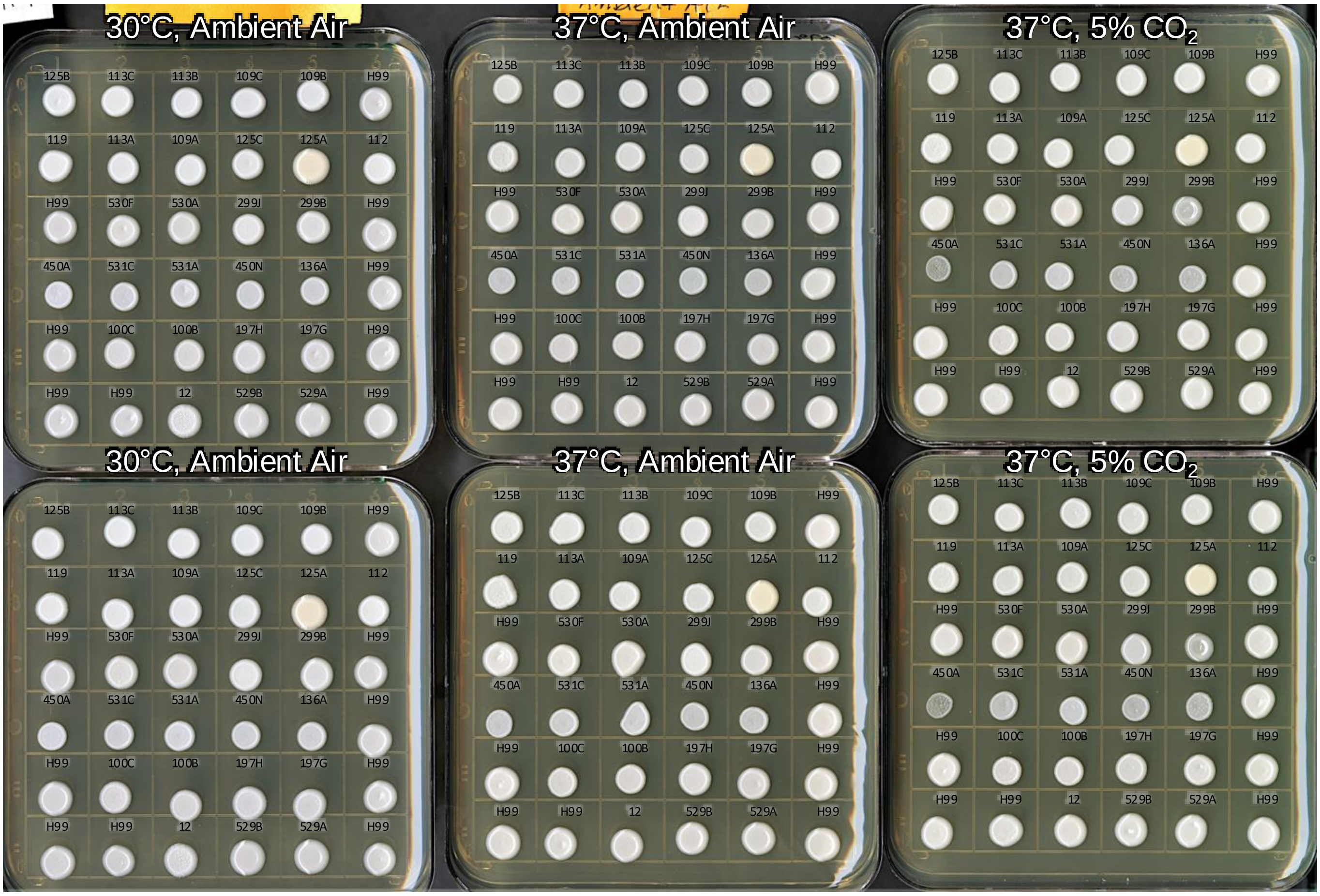


**Figure S4 Continued. Phenotypic variation and stress-dependent phenotypic switching on YPD agar after 72 hours of incubation is observed among the colonies isolated from serial isolates from patients NC8, NC83, NC94, NC183, and NC6. E)** Differences in colony complexity is observed between the colonies isolated from serial specimens of each patient. Phenotypic switching with regard to colony complexity is also observed among isolates from at least one colony from each patient. Top two rows are patient NC8 strains, the third row NC83, fourth row NC94, fifth row NC183, and the bottom row is NC6 strains. CTRL – control strain H99. MCM125A was identified to be *Cryptococcus albidus* indicating that the CSF specimen MCM125 represented a timepoint in which patient NC8 had a mixed-cryptococcal species infection.

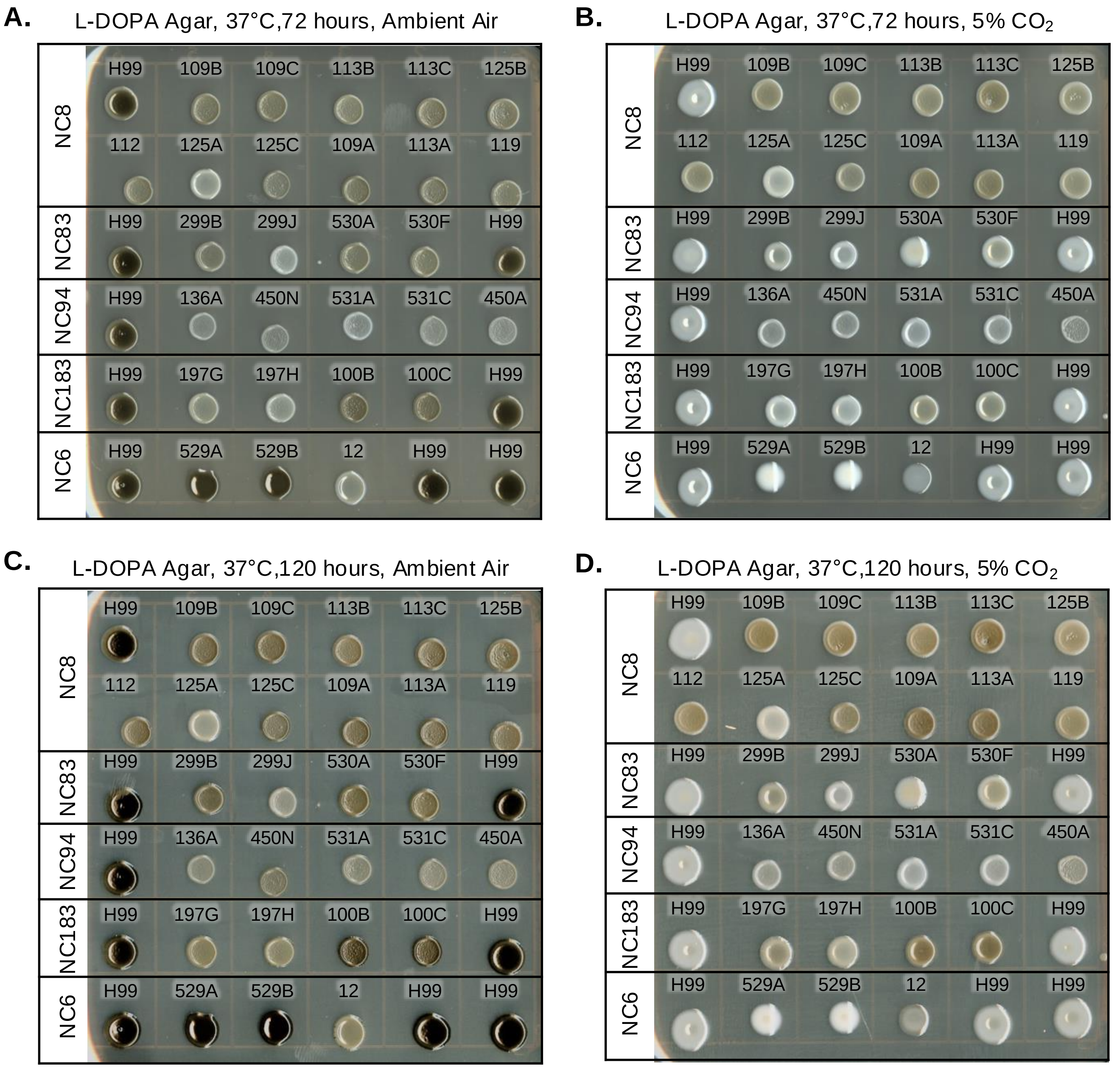


**Figure S5. Phenotypic variation and stress-dependent phenotypic switching on melanin-inducing L-DOPA agar is observed among the colonies isolated from serial isolates from patients NC8, NC83, NC94, NC183, and NC6. A-D)** Differences in colony pigment is observed between the colonies isolated from serial specimens of each patient after incubating for 72 hours and 120 hours. Phenotypic switching with regard to colony viscosity and melanin pigment is also observed among isolates from at least one colony from each patient. $CO_2$-dependent delay in melanin formation is observed up to 120 hours of incubation for some strains. Patient NC8's strains did not display a marked $CO_2$-dependent delay in melanin formation. MCM125A was identified to be *Cryptococcus albidus* indicating that the CSF specimen MCM125 represented a timepoint in which patient NC8 had a mixed-cryptococcal species infection. was used as a control strain.

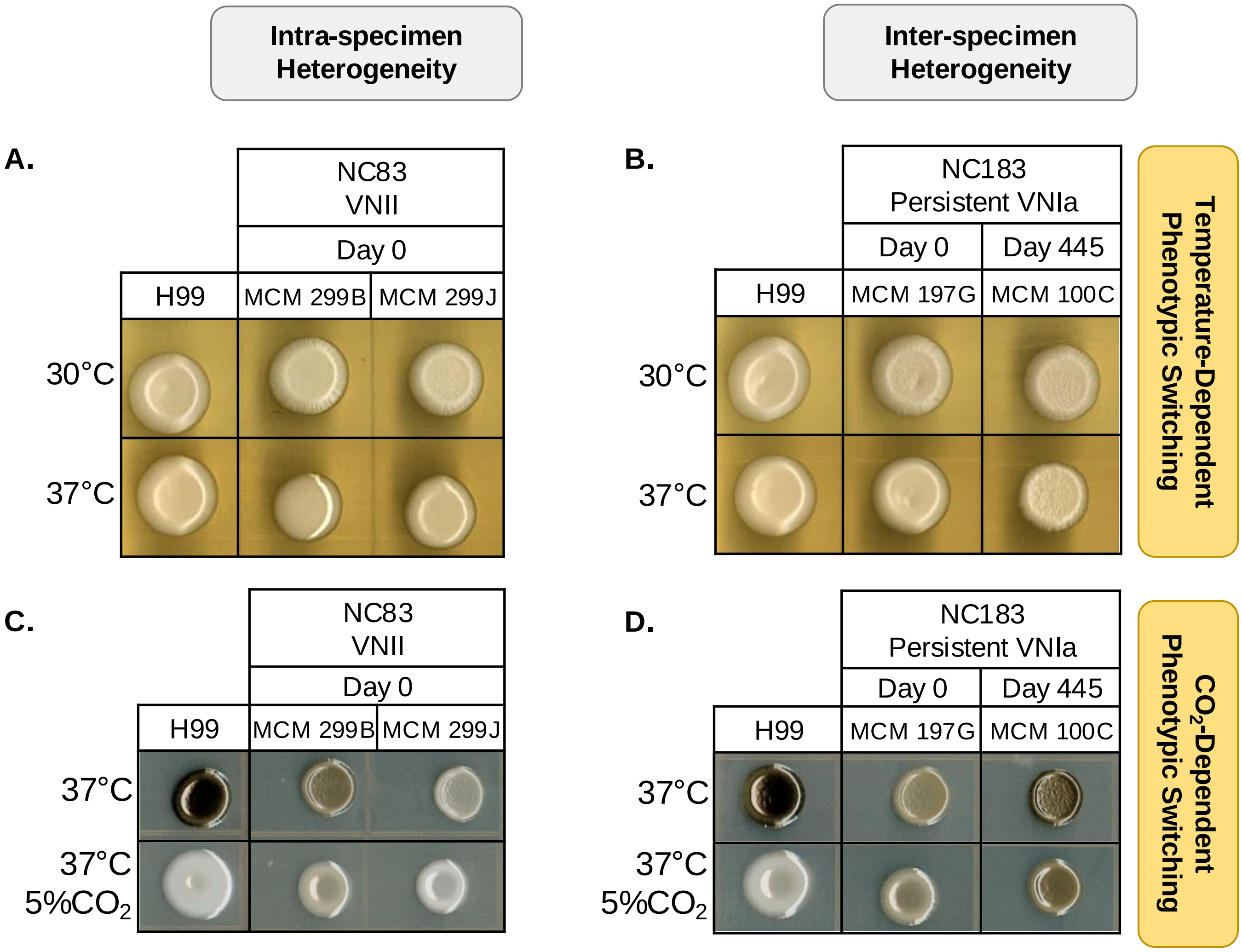


**Figure S6. Phenotypic heterogeneity and stress-dependent phenotypic switching of colony complexity, viscosity, and melanin pigment are visualized within individual patient specimens (intra-specimen) and between serial specimens from the same patient (inter-specimen). A)** Intra-specimen phenotypic heterogeneity in viscosity occurs between two VNII colonies from patient NC83's incident specimen under high-temperature stress (37°C), indicating stress-dependent phenotypic switching in one colony but not the other at 30°C.**B)** Inter-specimen phenotypic heterogeneity and switching occur between VNIa colonies from patient NC183 at 30°C and 37°C over 445 days, with complexity changes observed in colony morphology and stress-dependent phenotypic switching. **C)** Intra-specimen heterogeneity in melanin pigment is observed in two VNII colonies from patient NC83 under high-temperature stress, with no growth impact, an exacerbated stress-dependent delay in melanin formation, and a switching of colony viscosity. **D)** Inter-specimen phenotypic heterogeneity is evident in patient NC183's specimens, with strain MCM197G (day 0) lighter than MCM100C (day 445), showing stress-dependent delay in melanin formation and phenotypic switching from creamy to mucoid.

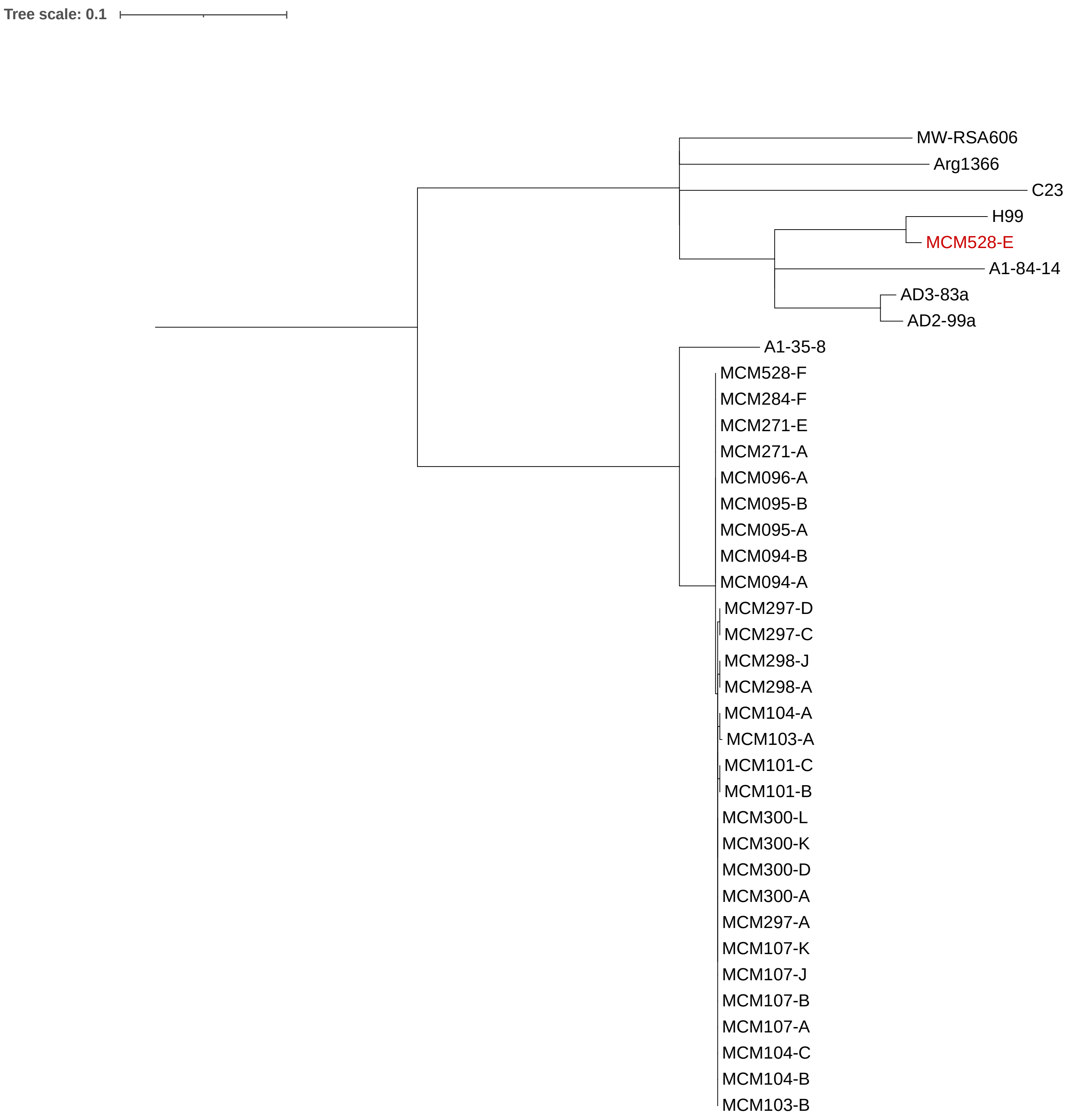


**Figure S7. Maximum-likelihood tree for strains isolated from patient NC1.**
A maximum-likelihood tree for isolates from patient NC1 (MCM strain IDs) and other strains closely related to the reference strain H99. Each strain was mapped to the genome of H99, and an alignment of SNP sites was used as the input to the program RAxML-NG.

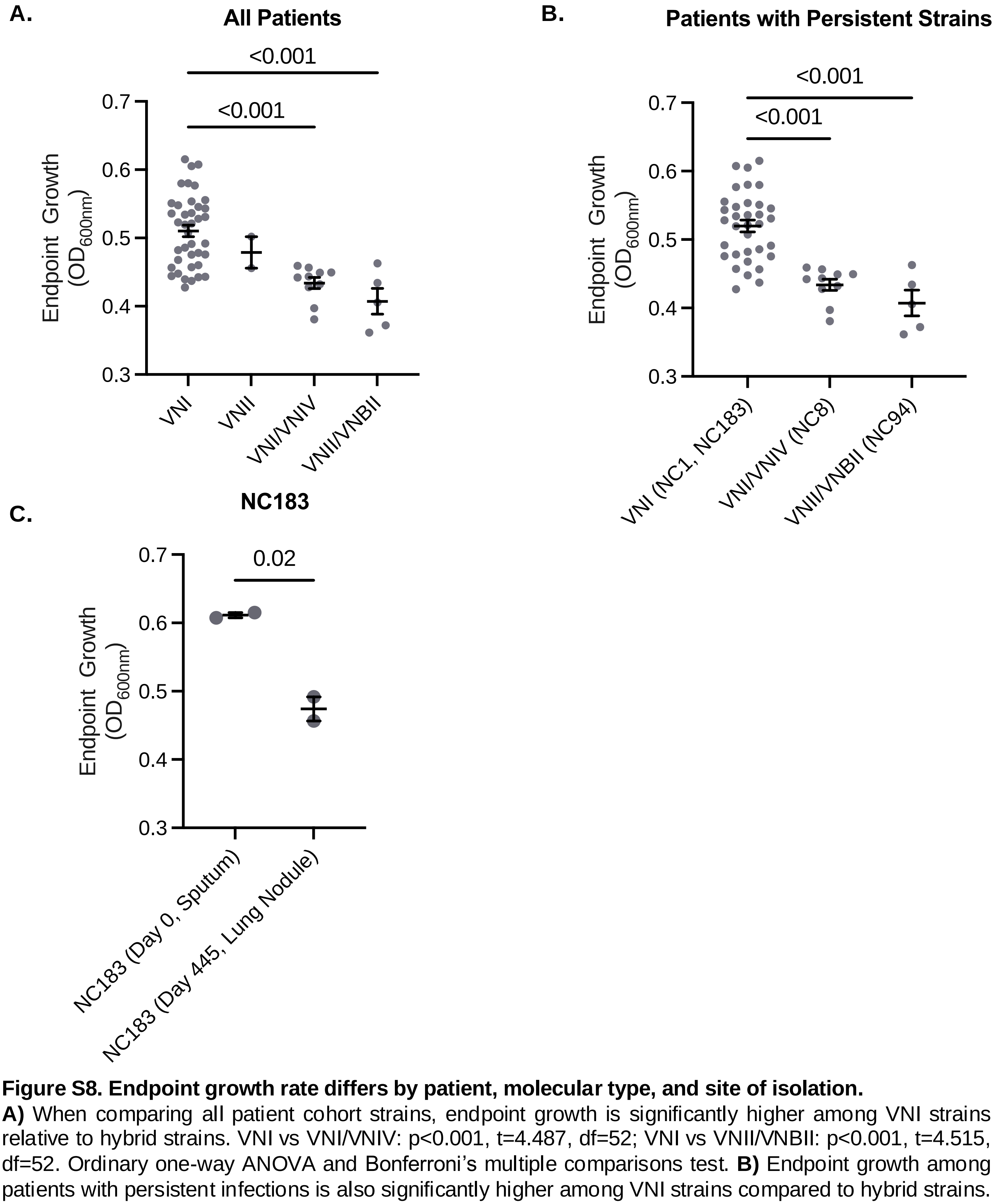


**Figure S8. Endpoint growth rate differs by patient, molecular type, and site of isolation.**
**A)** When comparing all patient cohort strains, endpoint growth is significantly higher among VNI strains relative to hybrid strains. VNI vs VNI/VNIV: p<0.001, t=4.487, df=52; VNI vs VNII/VNBII: p<0.001, t=4.515, df=52. Ordinary one-way ANOVA and Bonferroni's multiple comparisons test. **B)** Endpoint growth among patients with persistent infections is also significantly higher among VNI strains compared to hybrid strains. VNI/VNIV (NC8) vs. VNI (NC1, NC183): p<0.001, t=5.256, df=46; VNII/VNBII (NC94) vs. VNI (NC1, NC183): p<0.001, t=5.172, df=46. Ordinary one-way ANOVA and Bonferroni's multiple comparisons test. **C)** Patient NC183 had two strains identified in their sputum on day 0 and two on day 445 isolated from a lung nodule. The strains isolated from the sputum had a significantly higher endpoint growth rate compared to those isolated from the pulmonary nodule. p=0.02, t=7.585, df=2. Unpaired t-test.

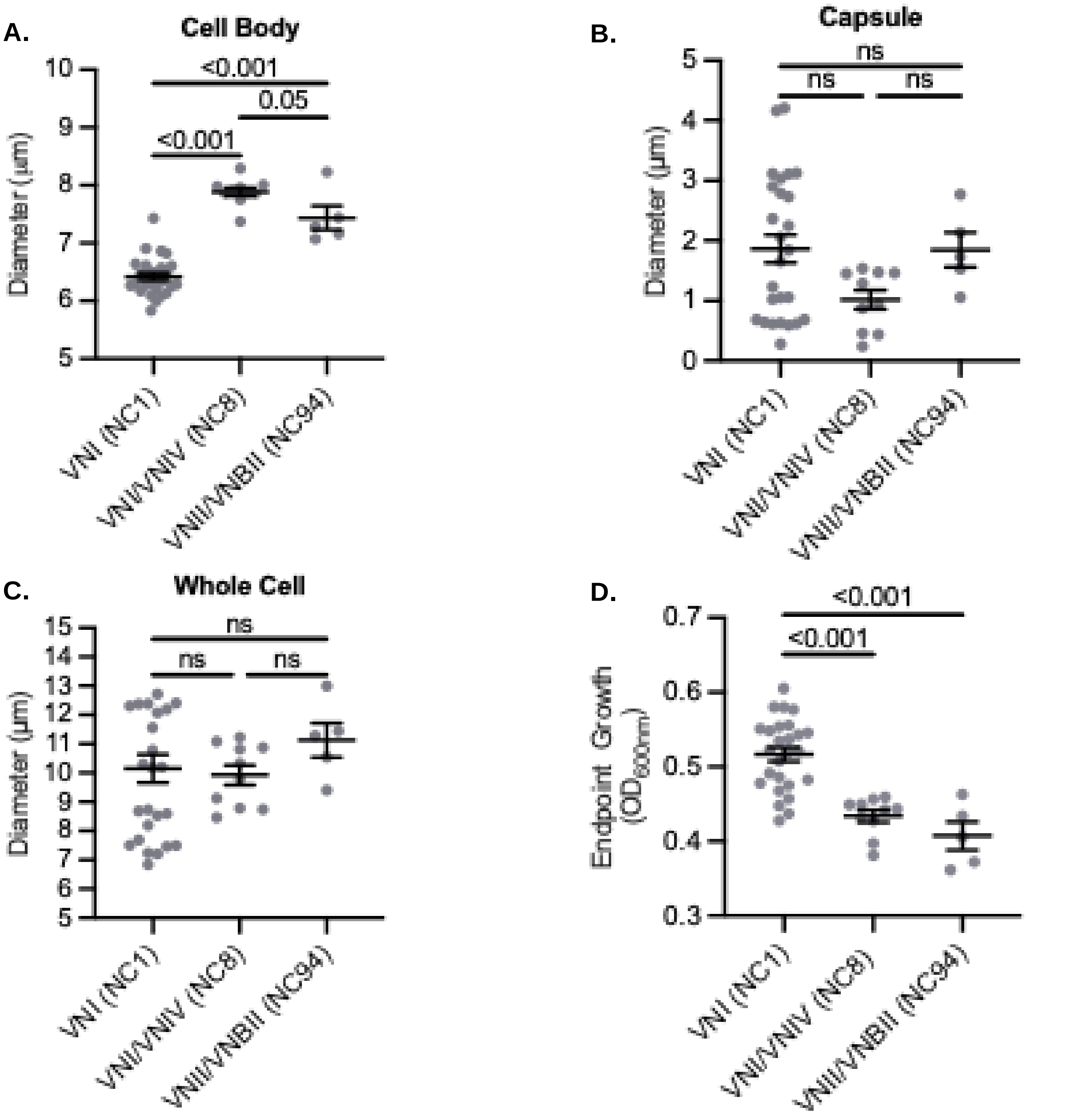


**Figure S9. Characterization of cell size attributes compared by patient and cryptococcal molecular type reveals distinct cell body sizes between groups and large variation of capsule within groups.**
**A)** Cell body diameter of strains collected from patient NC1 are significantly smaller than both hybrid strains. Also, the VNI/VNIV strains have a larger cell body diameter compared to both VNI and VNII/VNBII strains. VNI (NC1) vs. VNI/VNIV (NC8): p=<0.001 t=11.92 df=38; VNI (NC1) vs. VNII/VNBII (NC94): p=<0.001 t=6.303 df=38; VNI/VNIV (NC8) vs. VNII/VNBII (NC94): p=0.05 t=2.477 df=38. Ordinary one-way ANOVA and Bonferroni's multiple comparisons test. **B)** Capsule thickness of strains show greater variation within each group which results in no significant difference between groups. Kruskal-Wallis test and Dunn's multiple comparisons test. **C)** As a result of having little variation within each group for cell body size and large variation within each group for capsule thickness, there is no significant difference in whole cell diameter between groups. Kruskal-Wallis test and Dunn's multiple comparisons test. **D)** Despite having no significant difference in whole cell diameter, endpoint growth of VNI strains is significantly greater than both hybrid strains. Graph only contains data points from strains with cell size measurements. VNI (NC1) vs. VNI/VNIV (NC8): p=<0.001 t=5.151 df=38; VNI (NC1) vs. VNII/VNBII (NC94): p=<0.001 t=5.190 df=38. Ordinary one-way ANOVA and Bonferroni's multiple comparisons test.

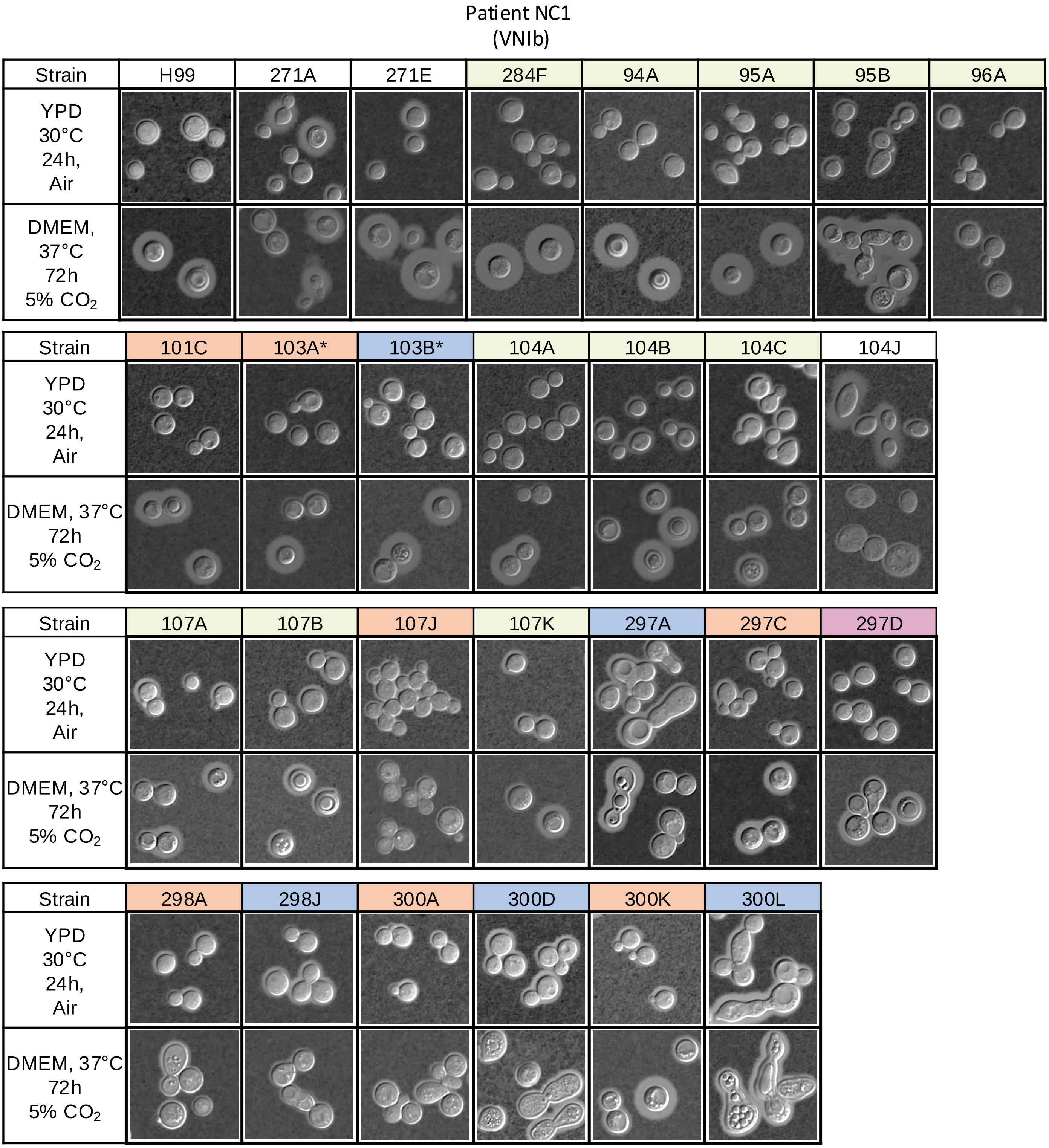


**Figure S10. Cell morphology of VNIb strains collected from patient NC1 under classical culturing conditions and host-like stress conditions.** Images are arranged in timeline order from top to bottom and left to right. Cell morphology changes over time along with specific genomic changes. White indicates euploid strains and green indicates strains with partial chromosome 10 deletion. The following indicate strains with partial chromosome 10 deletion in combination with a chromosomal duplication: orange color indicates strains with Chr.12 duplications, blue color represents strains with Chr.1 duplications, and magenta color represents strains with Chr.6 duplication. MCM103A and MCM103B each have partial duplications of Chr.12 and Chr.1 indicated by *, respectively. MCM104J was genotyped to be *Cryptococcus albidus*.

Patient NC1
(VNIb)

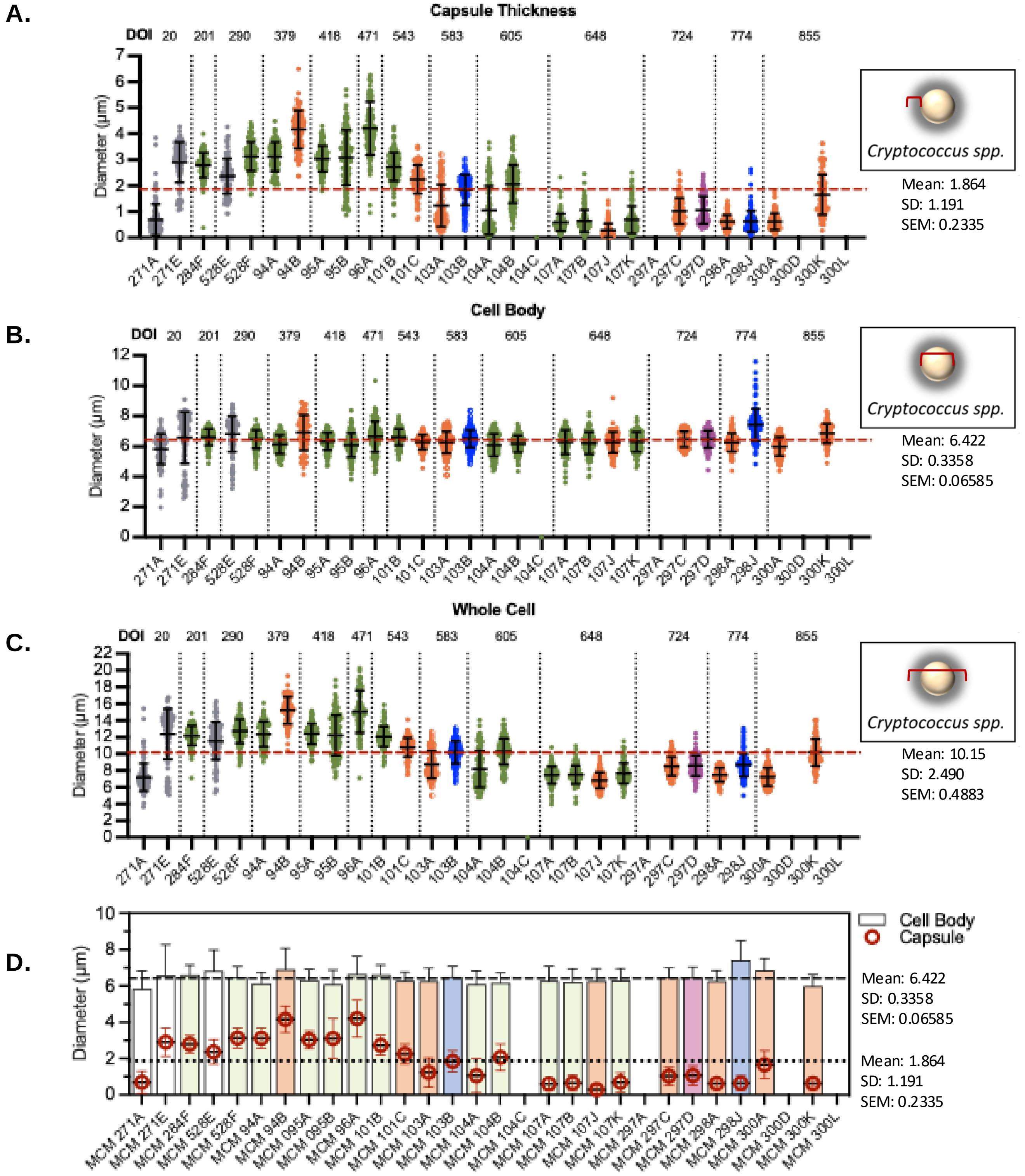


**Figure S11. Phenotypic characterization of cell morphology of VNIb strains collected from patient NC1 during persistent cryptococcal meningitis infection. A)** Diameter of capsule thickness shows a drastic decrease in size over time. **B)** Cell body diameter remains stable over time. **C)** Whole cell diameter also decreases over time as a result of the decrease in capsule thickness. **D)** Overlay of cell body diameter and capsule thickness diameter shows change over time. Grey or white indicates euploid strains and green indicates strains with partial chromosome 10 deletion. The following indicate strains with partial chromosome 10 deletion in combination with a chromosomal duplication: orange indicates strains with Chr12 duplication, blue indicates strains with Chr1 duplication, magenta indicates strains with Chr6 duplication.

Patient NC1
(VNIb)

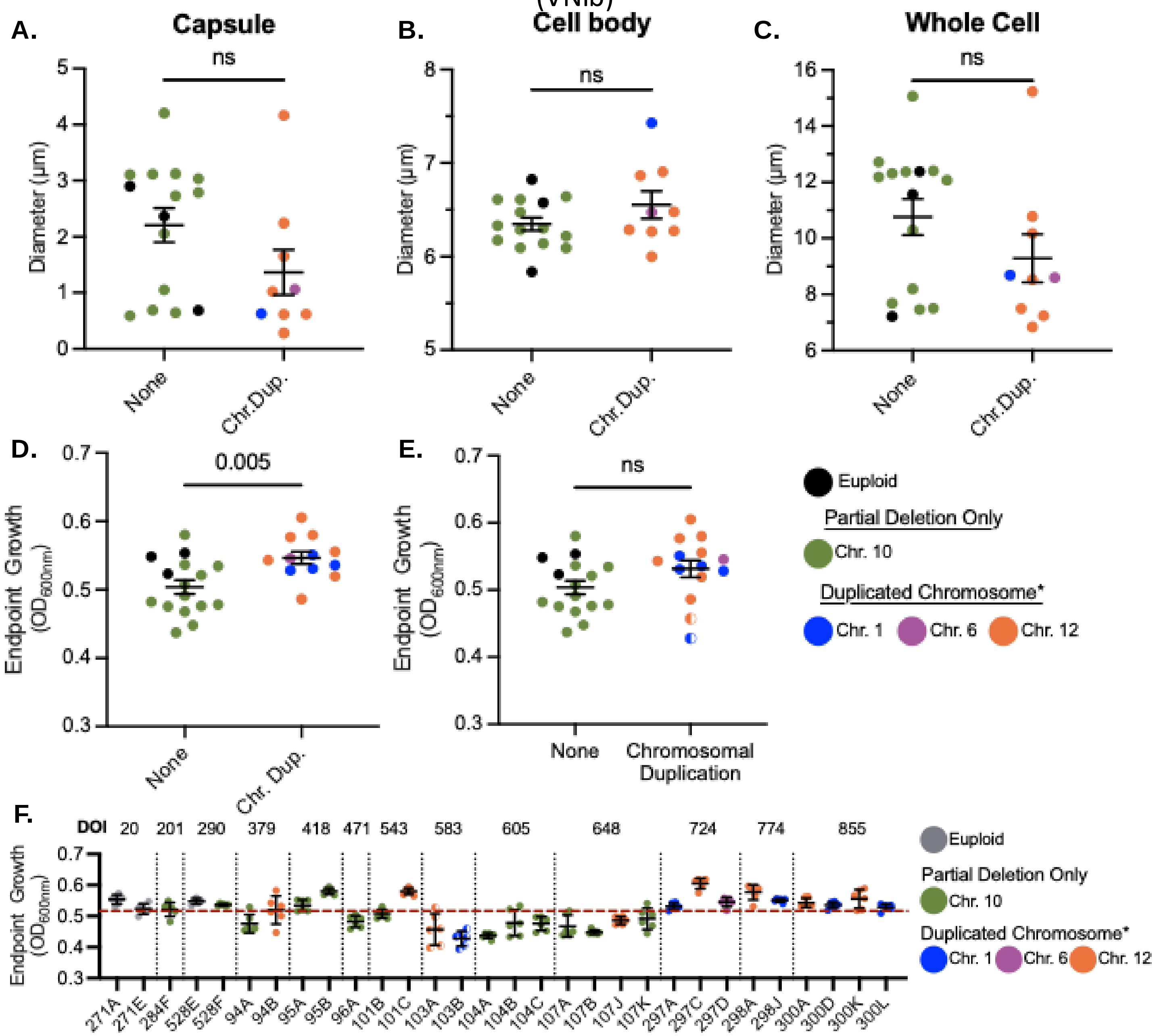


**Figure S12. Patient NC1 VNIb strains with chromosomal duplications have greater endpoint growth but smaller whole cell size due to smaller capsule thickness.** Bars and values below represent mean and standard error of the mean. Significance calculated using unpaired t-test. **A-C)** There is no significant difference between capsule, cell body, or whole cell diameter when comparing strains with or without large chromosomal duplications. **D)** Endpoint growth of VNIb strains with large chromosomal duplications have significantly greater endpoint growth compared to strains without large genomic changes. None mean= 0.5038 ± 0.01013, Chr.Dup. mean=0.5464 ± 0.008993, p=0.005, t=3.030, df=26. Strains included in endpoint growth are those with large chromosomal duplications only (excludes MCM103A and MCM103B). **E)** Endpoint growth, when including strains that had both large chromosomal duplications and those with partial chromosome duplications (MCM103A and MCM103B) shows no significant difference between strains without chromosomal duplications and those with duplications. None mean= 0.5038 ± 0.01013, Chromosomal Duplication mean=0.5315 ± 0.01276, p=0.10, t=1.724, df=28. F) Overview of endpoint growth for all strains with a dashed line represents mean =0.5167 ± 0.008306 n=30. *Strains with chromosomal duplications also harbor a partial Chr.10 deletion. Half filled circles represent partial duplication.

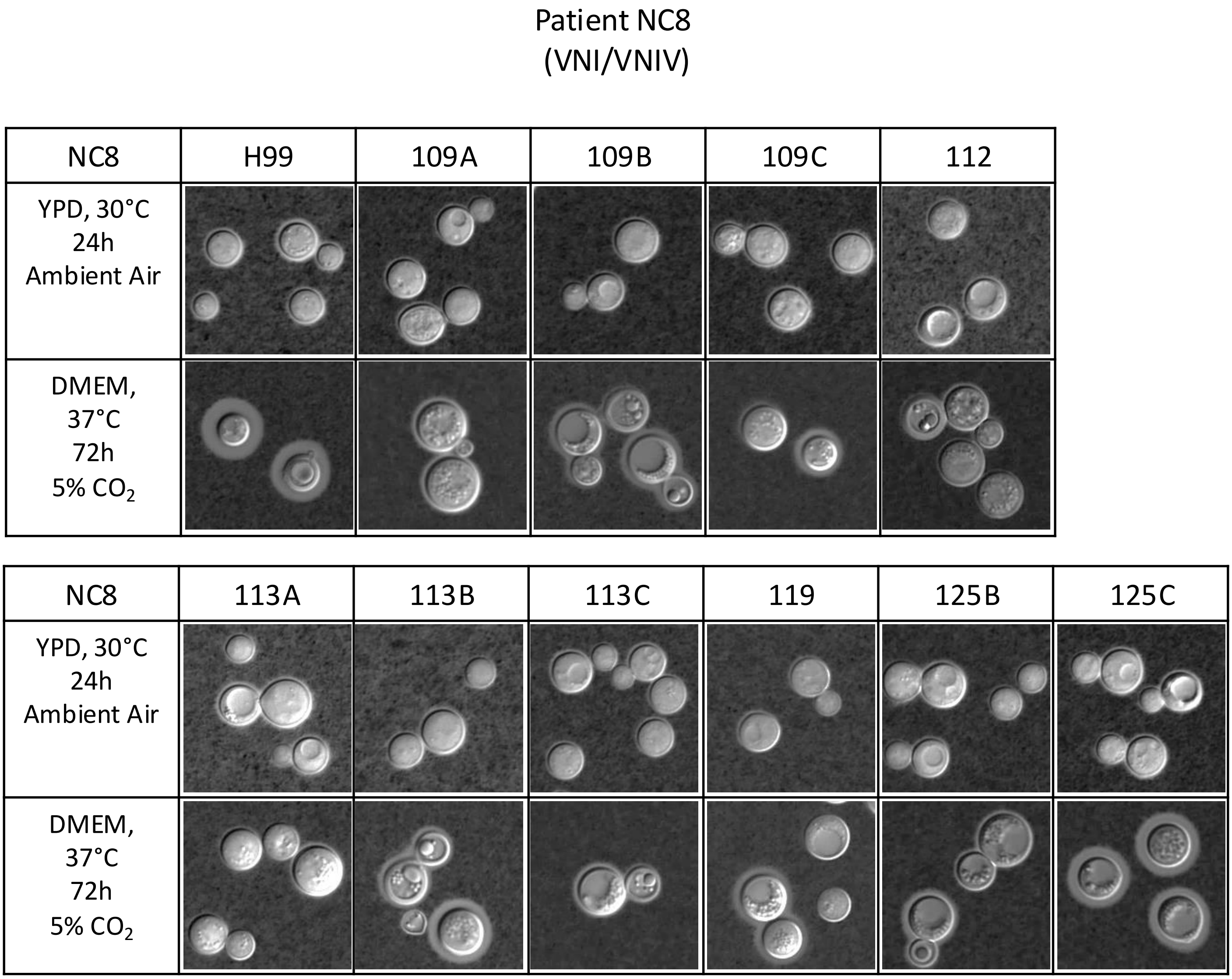


**Figure S13. Cell morphology of VNI/VNIV hybrid strains collected from patient NC8 under classical culturing conditions and host-like stress conditions.** Images are arranged in timeline order from top to bottom and left to right. Cell morphology does not show significant changes over time.

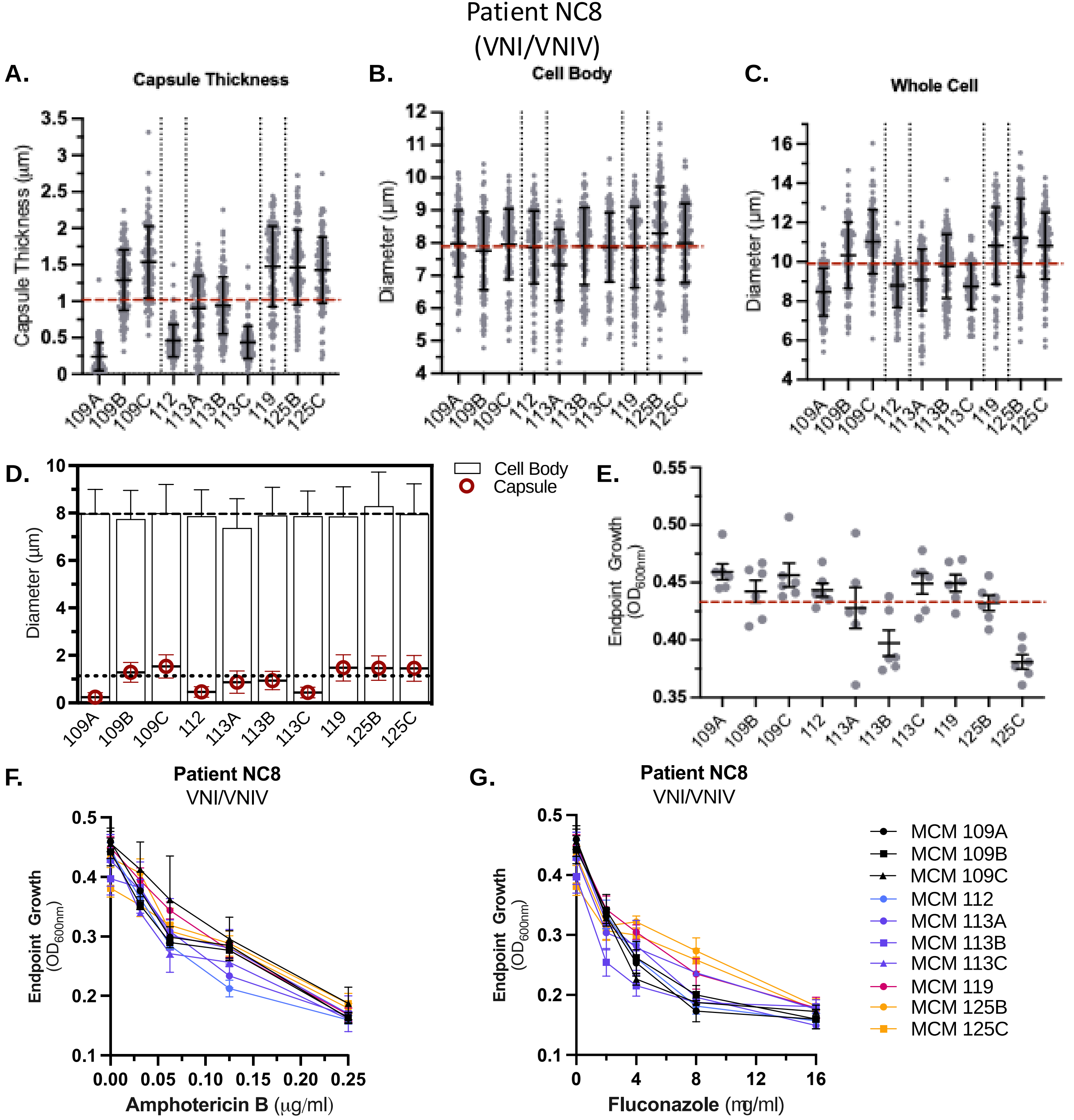


**Figure S14. Phenotypic characterization and antifungal susceptibility of VNI/VNIV hybrid strains collected from patient NC8 during persistent cryptococcal meningitis infection.** A-D) Bars represent mean and standard deviation, E-G) Bars represent mean standard error of the mean. **A)** Capsule thickness shows variation over time. Red line is the average capsule thickness of all isolates (Avg = 1.018 ± SD = 0.4969, SEM = 0.1571). **B)** Cell body diameter remains relatively similar throughout all timepoints. Red line is the average cell body diameter of all isolates (Avg = 7.887 ± SD = 0.2292, SEM = 0.07248). **C)** Variation in whole cell diameter is a result of the variation in capsule thickness among the strains. Red line is the average capsule thickness of all isolates (Avg = 9.923 ± SD = 1.075, SEM = 0.3398). **D)** Another representation of how cell body and capsule diameters change over time for each strain. **E)** Red line is the average endpoint growth $OD_{600nm}$= 0.4339 ± SD = 0.02576, SEM = 0.008145. **F)** Amphotericin B antifungal susceptibility measured by endpoint growth. **G)** Fluconazole antifungal susceptibility measured by endpoint growth.

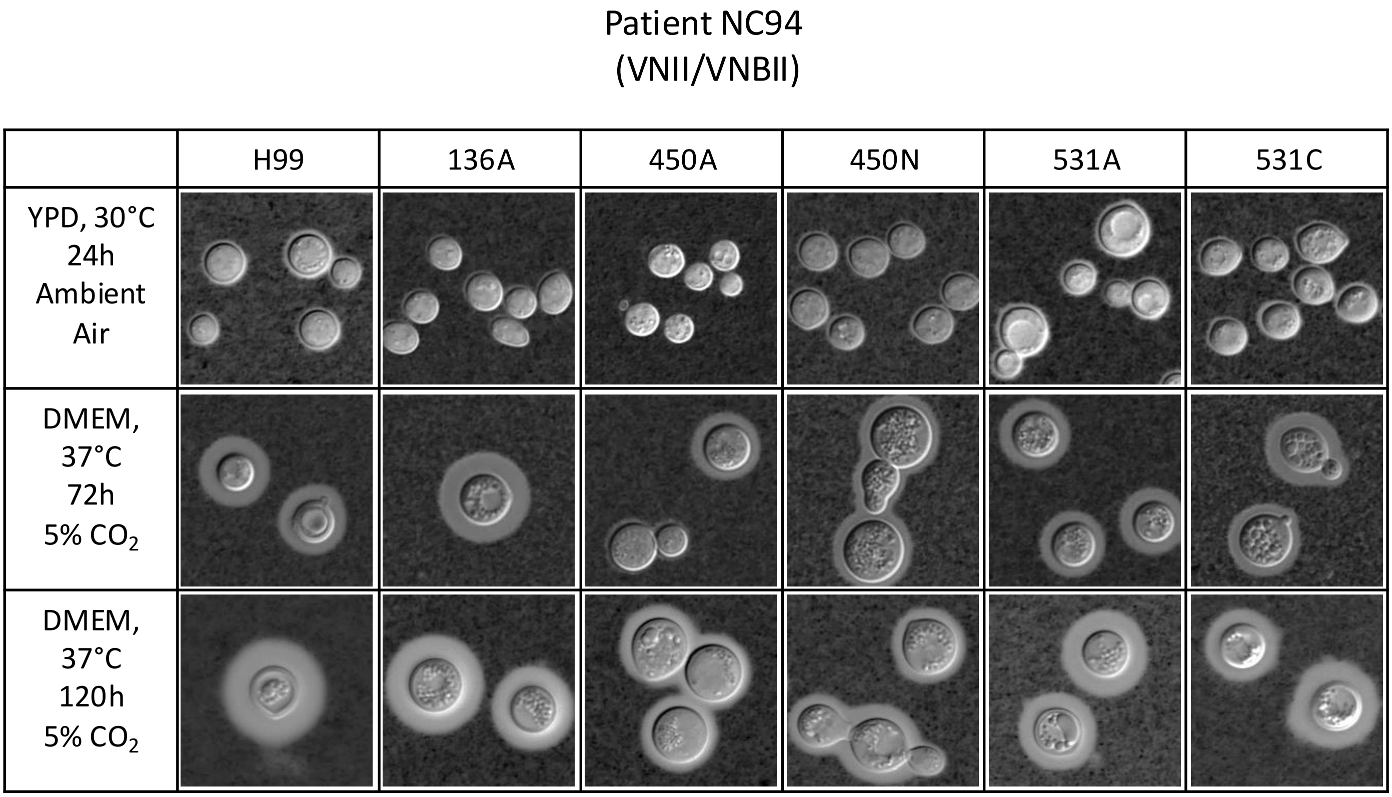


**Figure S15. Cell morphology of VNII/VNBII hybrid strains collected from patient NC94 under classical culturing conditions and host-like stress conditions.** Images are arranged in timeline order from top to bottom and left to right. Cell morphology differs between strains. Specifically, MCM450A produces cells that are either round or teardrop shaped under host-like stress conditions. MCM450N also produces cells with either round or teardrop shape along with a seemingly cell separation defect or pseudohyphae. MCM531C also produces round or teardrop shaped cells.

Patient NC94
(VNII/VNBII)

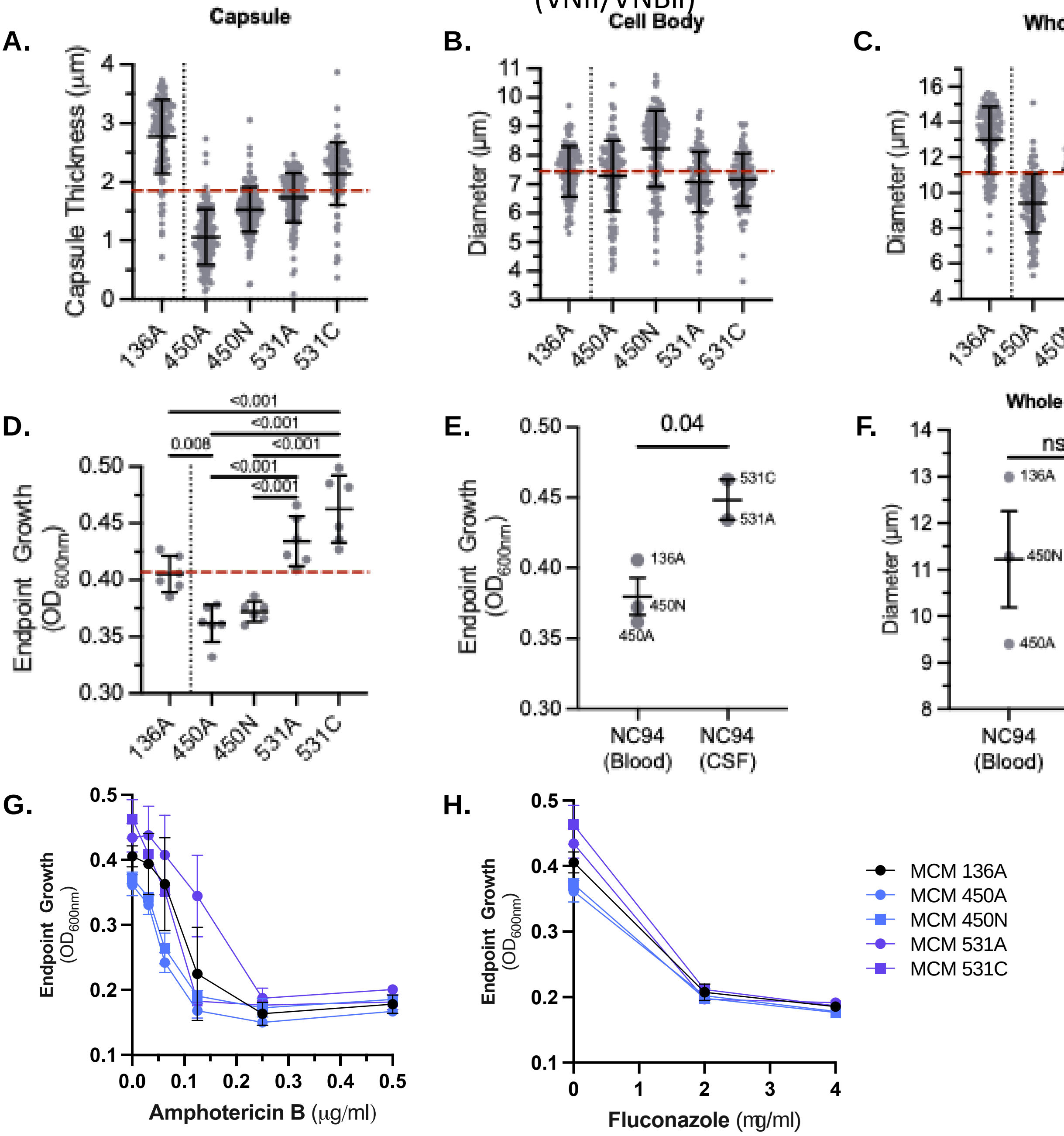


**Figure S16. Phenotypic characterization and antifungal susceptibility of VNII/VNBII hybrid strains collected from patient NC94 during persistent cryptococcal meningitis infection. .** A-D) Bars represent mean and standard deviation, E-H) Bars represent mean standard error of the mean. **A)** Capsule thickness shows variation over time. Red line is the average capsule thickness of all isolates (Avg = 1.846 ± 0.2896). **B)** Cell body diameter remains relatively similar throughout all timepoints. Red line is the average cell body diameter of all isolates (Avg = 7.439 ± 0.2069 ). **C)** Variation in whole cell diameter is a result of the variation in capsule thickness and cell body size among the strains. Red line is the average capsule thickness of all isolates (Avg = 11.13 ± 0.5876). **D)** Endpoint growth of strains shows a significant change over time and by body site. Red line is the average endpoint growth $OD_{600nm}$= 0.4072 ± SEM = 0.01885. Individual statistics can be found in supplementary data files. **E)** Endpoint growth of strains shows a significant change by body site. NC94(Blood) Avg. = 0.3798 ± SEM = 0.01321; NC94(CSF) Avg. = 0.4484 ± SEM = 0.01425; p= 0.04, t= 3.416, df = 3. **F)** There is no significant difference in whole cell size by body site. This suggests CSF isolates grow faster than the blood isolates. **G)** Amphotericin B antifungal susceptibility measured by endpoint growth. **H)** Fluconazole antifungal susceptibility measured by endpoint growth.

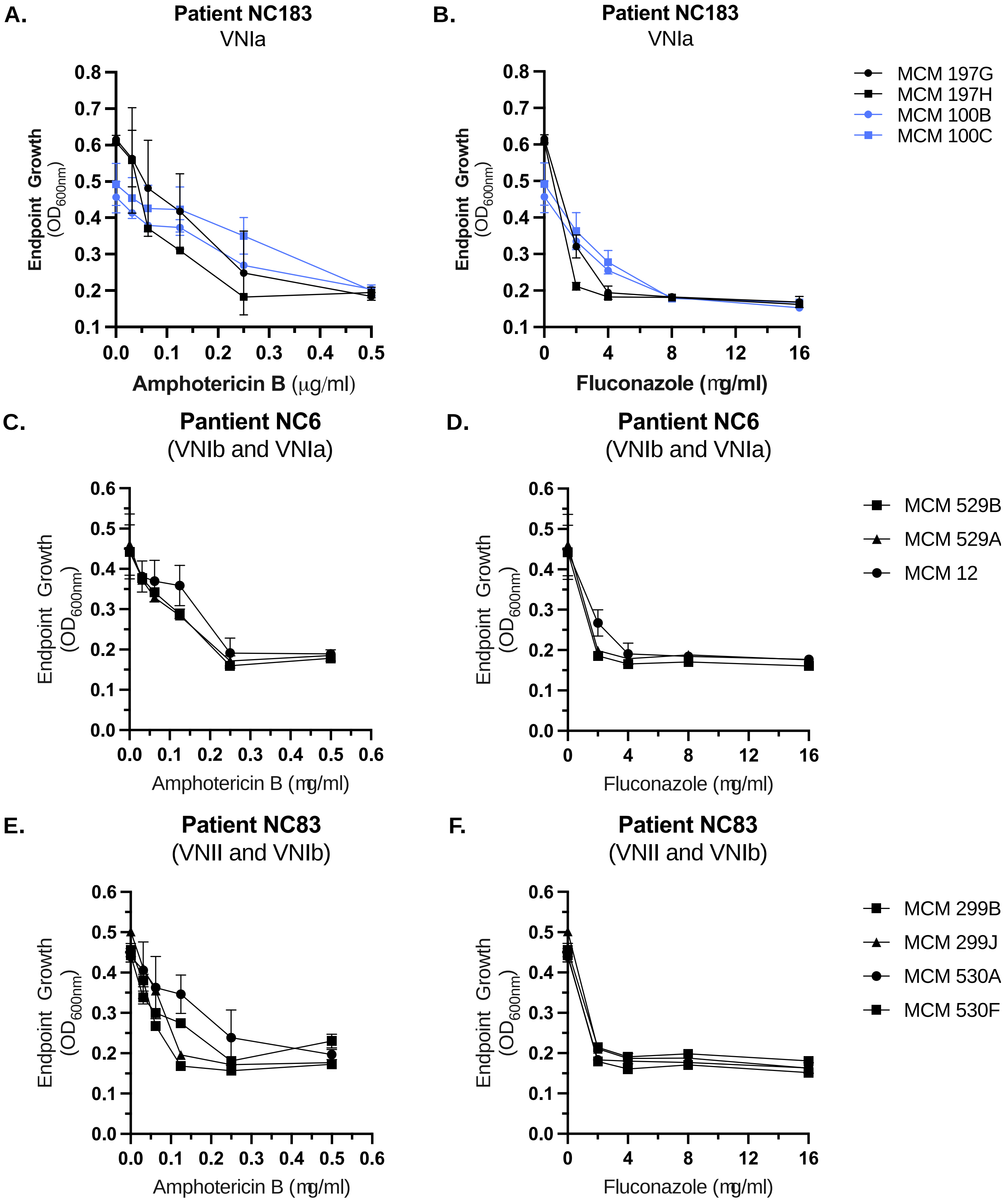


**Figure S17. Antifungal susceptibility assessed through optical density measurements of strains from cryptococcosis patients with novel strain reinfections.**

**Patient NC1 (VNIb)**

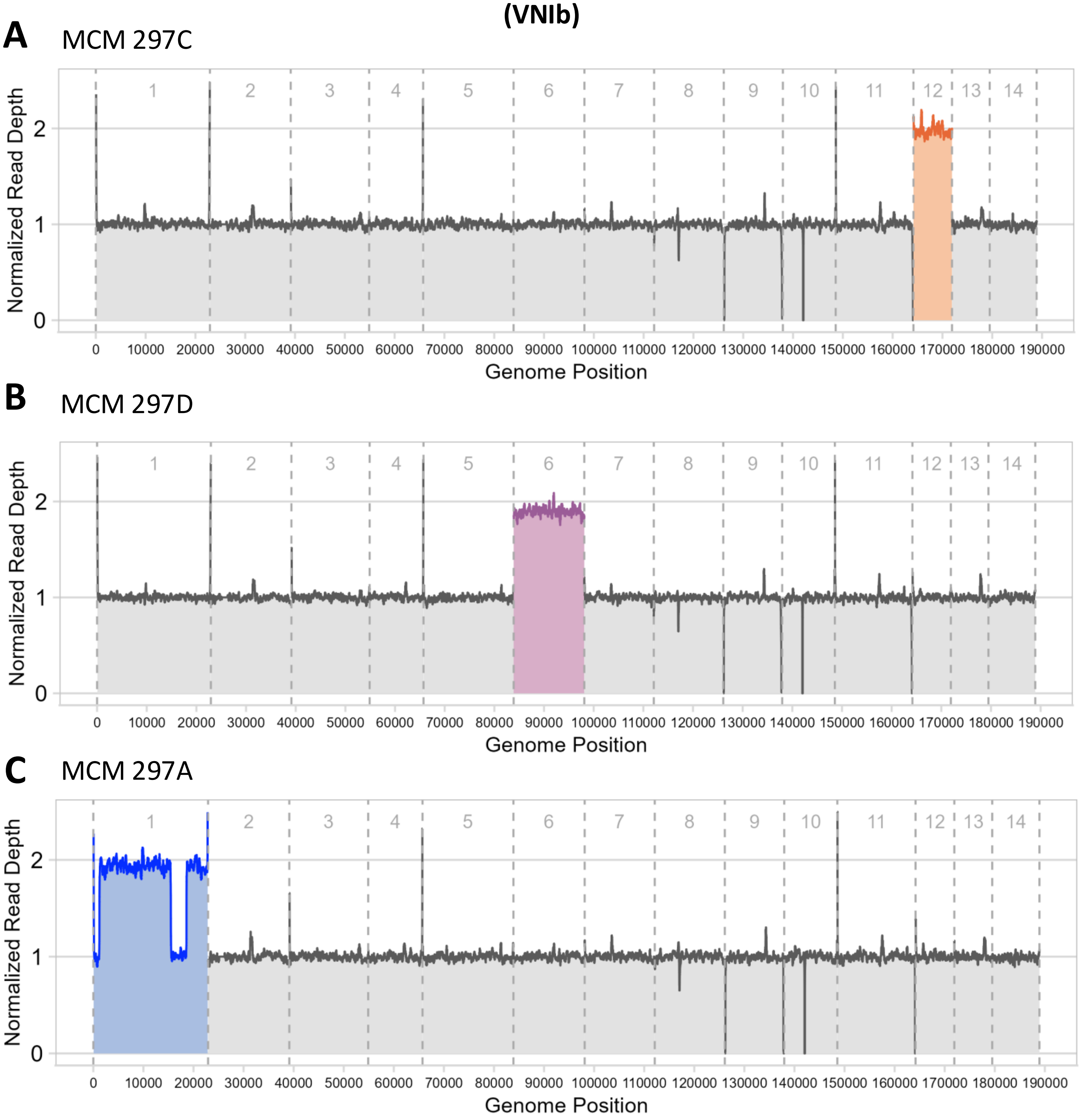


**Figure S18. Chromosomal duplications in NC1 lineages assessed through normalized read depth.** Genomic read depth profiles of three different NC1 strains show full-chromosome duplications of chromosome 12 **(A)** and chromosome 6 **(B)** and a partial duplication of chromosome 1 **(C).** Read depth is measured by alignment to the H99 reference genome and normalized to the median within each sample. Normalized read depth of 1 represents a euploid chromosome, while a normalized read depth of 2 represents a duplicated region. The entire genome is shown with the chromosomes segmented and numbered from left to right.

**Patient NC1**
**(VNIb)**

Chromosome 1

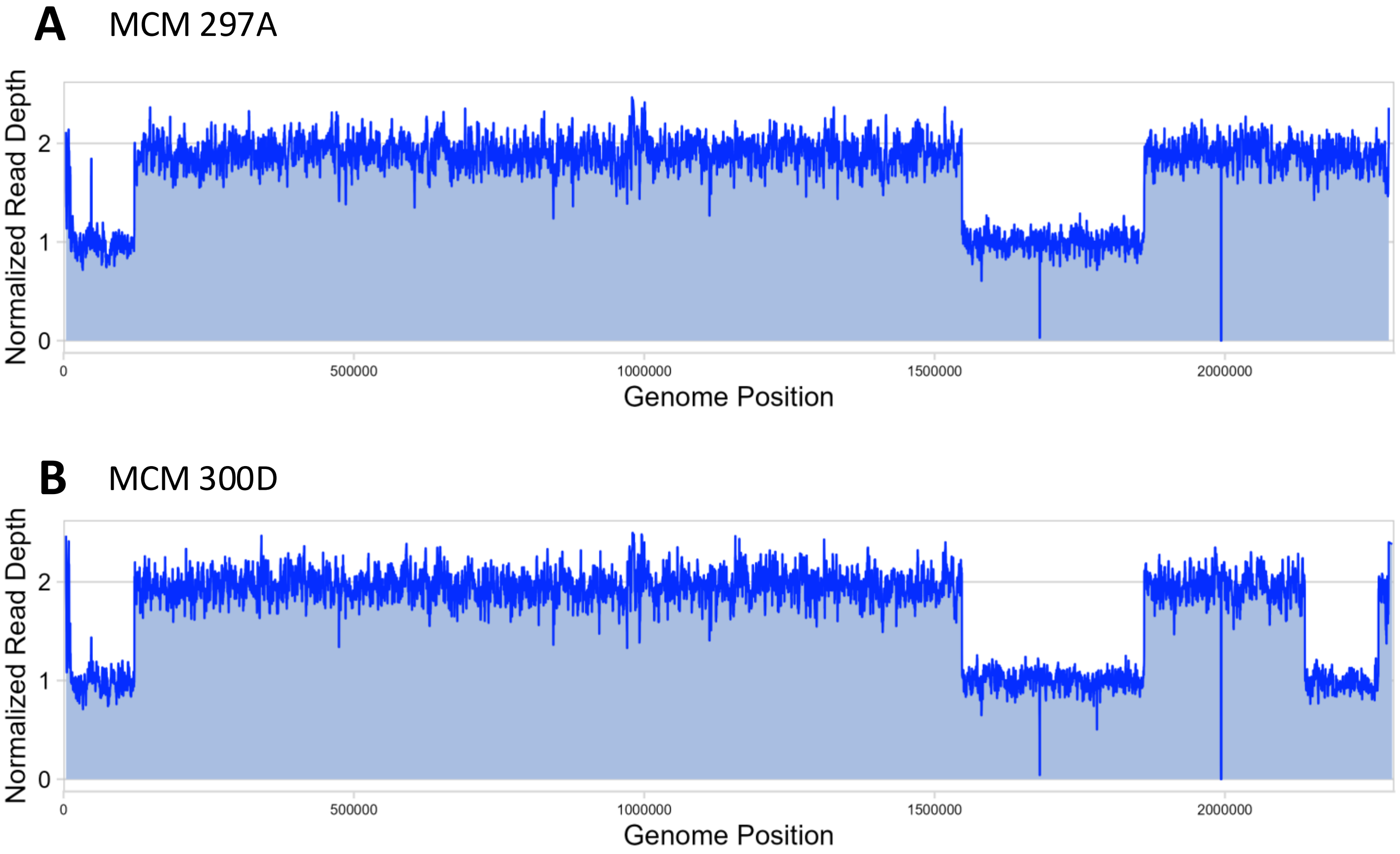


**Figure S19. Loss of a chromosome 1 duplicated region in NC1 lineages assessed through normalized read depth.** Read depth analysis of the partially duplicated chromosome 1 lineage in patient NC1 reveals a region on the right arm that is originally duplicated at an earlier timepoints of DOI 724 and DOI 774 **(A)** reverts back to euploid coverage at the last timepoint of DOI 855 **(B).** The newly-euploid region spans between the coordinates 2137500 – 2264500, while the originally euploid regions are located between 2250 – 122351 and 1546850 – 1861201 respectively. Read depth is measured by alignment to the H99 reference genome and normalized to the median within each sample. Normalized read depth of 1 represents a euploid chromosome, while a normalized read depth of 2 represents a duplicated region.
